# Physics-Based Growth and Remodeling Modeling for Virtual Abdominal Aortic Aneurysm Evolution and Growth Prediction

**DOI:** 10.64898/2026.02.26.26347026

**Authors:** Faeze Jahani, Zhenxiang Jiang, Malikeh Nabaei, Seungik Baek

## Abstract

Computational growth and remodeling (G&R) models have been extentively used to investigate abdominal aortic aneurysm (AAA) progression and to support clinical decision-making. However, the development of robust predictive models is often limited by the scarcity of large-scale longitudinal imaging datasets. In this study, we propose a physics-based G&R framework to simulate AAA shape evolution and generate a virtual cohort of aneurysms, thereby addressing data limitations and enabling integration with data-driven machine learning approaches for growth prediction. The proposed arterial G&R model incorporates key mechanisms influencing aneurysm progression, including elastin degradation and stress-mediated collagen production. A modified elastin degradation formulation was introduced to generate realistic aneurysm geometries exhibiting clinically relevant features such as asymmetry and tortuosity. By systematically varying parameters governing elastin damage and collagen production, 200 distinct G&R simulations were performed to produce a diverse set of AAA geometries. The dataset was further expanded using kriging-based spatial interpolation to construct a large in silico cohort. The synthetic dataset, combined with longitudinal imaging data from 25 patients, was used to train and validate four machine learning models: Deep Belief Network (DBN), Recurrent Neural Network (RNN), Long Short-Term Memory (LSTM), and Gated Recurrent Unit (GRU). A two-step training strategy was adopted to predict maximum aneurysm diameter and growth rate based on prior geometric characteristics. The LSTM model achieved the highest performance for maximum diameter prediction (R² = 0.92), while the RNN demonstrated strong overall performance (R² = 0.90 for maximum diameter and 0.89 for growth rate). The DBN and GRU models also showed competitive predictive capability. Overall, this study demonstrates that integrating physics-based G&R simulations with machine learning enables accurate prediction of AAA growth and maximum diameter. The proposed framework provides a scalable strategy for augmenting limited clinical datasets and offers a promising tool to support personalized risk assessment and treatment planning.

## 1. Introduction

An abdominal aortic aneurysm (AAA) is an abnormal localized dilation that occurs below the renal arteries, characterized by a maximum diameter exceeding 3 cm; small AAAs are typically monitored using imaging techniques such as ultrasound or CT scan. When an aneurysm enlarges to about 5–5.5 cm, surgical repair is usually advised, taking into account individual patient risk profiles [1]. An accurate prediction of AAA expansion is particularly valuable for improving clinical and surgical decision-making. Artificial intelligence (AI) and machine learning (ML) show promise for enhancing predictive accuracy in AAA risk assessment and growth [2–5], though access to follow-up imaging datasets remains limited. Consequently, synthetic or virtual image datasets have become alternative resources for training predictive algorithms [6,7]. Beyond physics-based virtual cohorts, alternative strategies have emerged to capture AAA shape evolution from sparse longitudinal data [8]. Such approaches address data scarcity from a complementary perspective, focusing on smooth fitting of observed shapes rather than simple geometrical training, and could potentially integrate with physics-based models for improved patient-specific predictions.

Nevertheless, the virtual dataset alone may not fully capture the complexities of aneurysm shape and biomechanical mechanisms of aneurysm growth in a real patient cohort. Recent work using hemodynamics-informatics has demonstrated that spatial patterns of wall shear stress and velocity can distinguish between fast- and slow-growing AAAs with high accuracy, highlighting the important role of local intraluminal flow in aneurysm progression [9]. Incorporating such hemodynamic signatures alongside computational modeling could provide additional predictive features and improve physiological realism in virtual cohort generation. Similar advances have been reported in thoracic aneurysms. Previous studies extracted local and global 3D shape features from longitudinal imaging of ascending aortas and showed that such morphological descriptors can predict growth more accurately than diameter alone. Other research showed that there is a direct link between growth and remodeling (G&R) model gain factors and AAA growth rate using early follow-up scans [10]. This supports the use of systematic geometry analysis as a complementary predictor in aneurysm growth modeling [11].

Meanwhile, substantial progress has been made in understanding vascular adaptation and disease progression [1]. In particular, the central roles of heterogeneous elastin degradation and ubiquitous stress-mediated G&R of collagen in AAA expansion have been increasingly revealed. Various physics-based computational G&R models have thus been developed on the basis of elastin damage and vascular adaptation to simulate AAA growth [12–17]. Those G&R models have been utilized to generate in silico datasets and then integrated with ML approaches to predict AAA shape evolution and growth rates [18–20]. Recent developments in geometric deep learning have shown that vascular shape changes can be predicted directly from clinical scans [21]. However, prior studies are often constrained by overly simplified damage functions and lack the ability to generate diverse and realistic geometries of AAAs.

To address this issue, this study introduces a novel elastin degradation function within a computational stress-mediated G&R model [20,22], enabling the generation of diverse and more realistic AAA geometries that account for asymmetry and tortuosity. Both characteristics have been linked to altered wall stress, accelerated aneurysm enlargement, and higher rupture risk, underscoring their importance in both mechanistic modeling and clinical decision-making [23,24] However, the asymmetric G&R simulation is computationally intense, necessitating the use of surrogate modeling to efficiently generate a high volume of datasets. Moreover, recognizing that clinical data alone is often insufficient, this study utilizes a dataset comprising follow-up CT images of AAAs from 25 patients [20] combined with in-silico data generated through the surrogate modeling based on the G&R model. These datasets are employed to train and test the ML models.

By integrating physics-based G&R simulation with data-driven and machine learning methods, we aim to overcome the challenges associated with limited clinical data and computational inefficiency, providing more accurate and efficient predictions of the AAA progression. Given that the maximum diameter and growth rate of AAA are crucial factors in clinical decision-making, ML models are specifically employed to predict these two parameters. The accuracy of different ML models in predicting the maximum diameter and growth rate was compared.

## 2. Methods

To provide a framework for predicting the growth and future behavior of AAAs, this study integrates a G&R computational model with machine learning tools through three main steps:

<u>G&R Computational Models</u>: A stress-mediated G&R model is employed to simulate the geometric evolution of AAAs. By specifying various elastin degradation functions and collagen remodeling rates, a wide range of realistic AAA geometries is generated.

<u>Surrogate Modeling for Data Generation</u>: The geometric data generated from the G&R simulations are further expanded using statistical models (kriging and the gradient-enhanced kriging with partial least squares (GEKPLS)), creating a comprehensive dataset for machine learning.

<u>Machine Learning for Predicting AAA Growth</u>: Various machine learning models, including Deep Belief Network (DBN), Recurrent Neural Network (RNN), Long Short-Term Memory (LSTM), and Gated Recurrent Unit (GRU), are trained using the data generated in the surrogate model. These models predict important features of AAAs, such as maximum diameter and growth rate.

### 2.1. G&R Simulation of the AAA

The central concept of the constrained mixture model (CMM) for soft tissue G&R was introduced by Humphrey and Rajagopal (2002) [25]. The framework is specifically designed to capture stress-mediated soft tissue adaptation over long time scales. In doing so, it neglects short-term thermodynamic dissipation (e.g., momentum exchange among solid constituents and biochemical reactions) and assumes a quasi-static equilibrium of linear momentum under mean physiological loading conditions (e.g., mean blood pressure). Then, vascular adaptation through G&R is modeled via the continual production and removal of constituents deposited with prescribed stretches (i.e., deposition stretches) with turnover rates governed by the current stress state (stress-mediated G&R). Over the past two decades, the constrained mixture framework has been further developed and extended, with its theoretical foundations and applications comprehensively reviewed by Humphrey (2021) [26]. Although the central principles of CMM are shared, specific formulations may vary among research groups [27–31].

The specific stress-mediated G&R formulation adopted for arterial tissue in this study is summarized in **Appendix**. Building on this theoretical framework, a computational model of AAA growth was developed to capture long-term changes in aneurysm geometry [20,25]. The model was implemented in membrane form using the finite element method (FEM) to simulate long-term shape evolution. The computational framework and the reduced elastin function are described in the following sections.

#### 2.1.1. Image-based G&R Computational Modeling Framework

In the presented CMM model, the initial elastin damage function represents a prescribed reduction in elastin content relative to a healthy vessel. As a consequence of the reduced elastin content, vessel wall stresses initially deviate from their homeostatic values, which in turn drives compensatory increases in other constituents (e.g., collagen and smooth muscle cells) to restore the target homeostatic stress state. The stress-mediated G&R formulation has been presented previously, and the relevant governing equations are provided in **Appendix A**. **Figure 1** illustrates the computational framework used to model AAA enlargement [32]. A three-dimensional geometric model is first constructed by segmenting medical images of a healthy aorta, from which a central region is selected for aneurysm simulations. Treating the vessel wall as a thin membrane, the surface is parameterized using longitudinal position along the arterial centerline and azimuthal orientation. Under normal physiological conditions, vascular constituents are produced and removed in balance, allowing the vessel to maintain a homeostatic configuration. In this framework, the image-based geometry is taken as the prestressed reference configuration. Because physiological pressure combined with spatial variations in material properties or wall thickness can perturb this configuration, an inverse optimization approach is employed to estimate distributions of material and geometric parameters (e.g., wall thickness and anisotropy) that preserve the in vivo geometry while satisfying target mechanical homeostasis [33,34]. More description of the optimization process and integral function of inverse optimization approach can be found in **Appendix B**.

**Figure 1.**
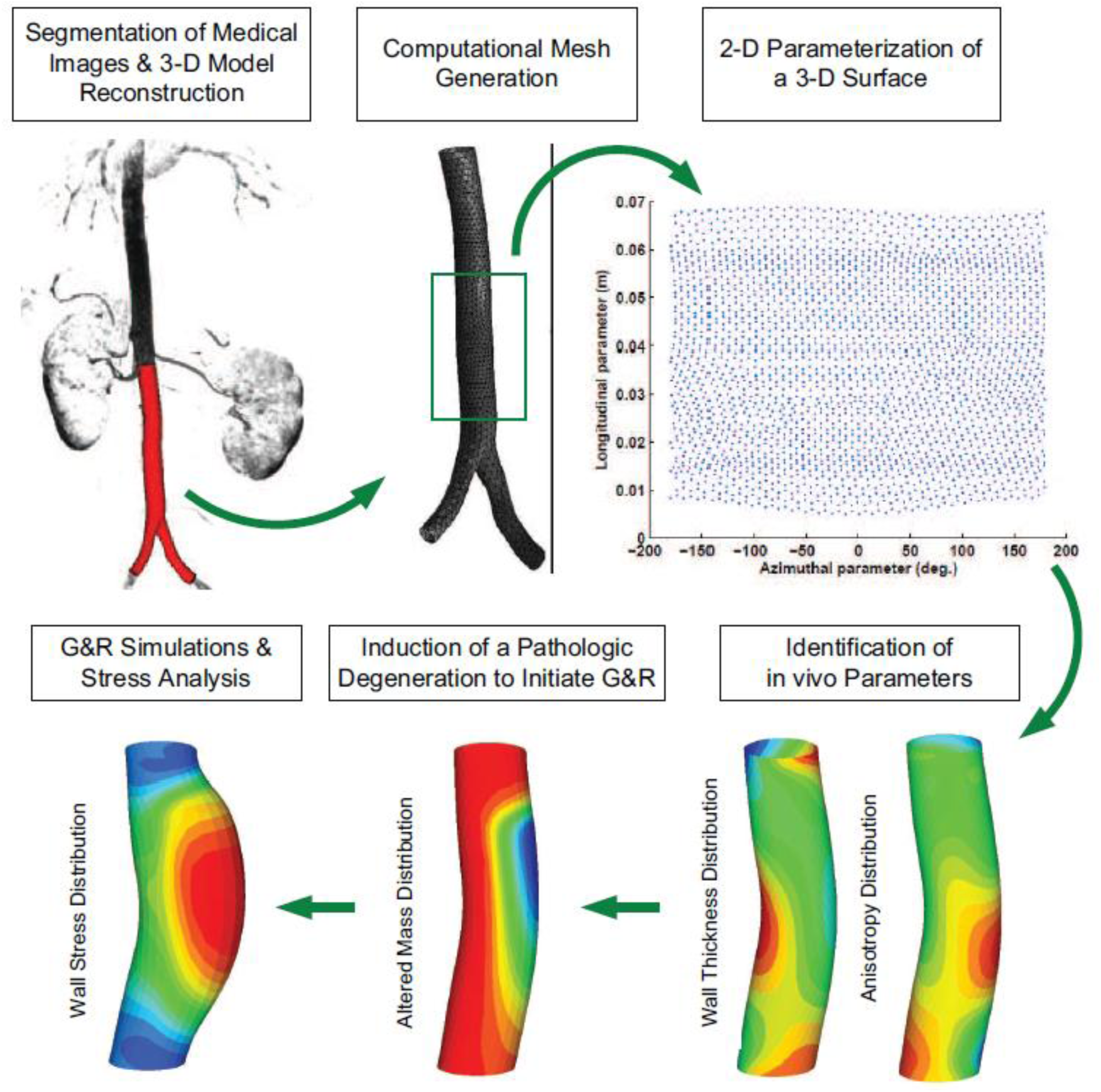
A schematic drawing of imaged-based G&R simulation[32].

#### 2.1.2. Modeling the Degradation of Elastin

Elastin degradation functions in previous studies [19,20] were only capable of producing symmetric growth or a bulged shape, in which the degradation variables were parameterized in a single axis, resulting in oversimplified AAA shapes. To cope with this problem and create geometries more consistent with medical images, a new initial elastin damage function was developed, incorporating a combination of two 2D Gaussian functions to model elastin degradation across different sections and circumferential locations of the cross-section.

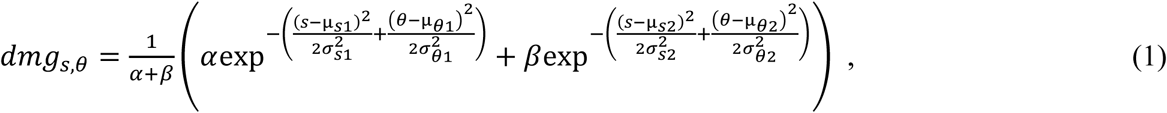

where *s* represents the length along the central axis direction and *θ* represents the angular coordinate in the circumferential direction; μ_*s*_ and μ_*θ*_ are used to determine the place where maximum elastin degradation occurs, μ_*s*_ identifies a point on the central axis where elastin degradation occurs, and μ_*θ*_ specifies the circumferential location of the degradation. In addition to determining the coordinates of the degradation, *σ*_*s*_ and *σ*_*θ*_ control the degradation area in the axial and circumferential directions. Furthermore, to normalize the damage function values between zero and one, parameters *ɑ* and *β* are utilized.

#### 2.1.3. Implementation in FEniCS and G&R Simulation Stages

The previous image-based FEM was originally developed using custom MATLAB code and has since been rewritten in FEniCS, an open-source computing platform for solving partial differential equations, to implement the G&R framework [20,35]. The corresponding FE formulation is provided in **Appendix C**. To ensure software compatibility and computational efficiency, all simulations and post-processing were performed in a containerized Linux environment using Docker.

The simulation of AAA growth involved two stages:

- Initial Simulation: This stage involved the first 300 days of achieving homeostatic equilibrium.
- Main Simulation: This stage simulated the growth of the AAA from day 300 to day 3600.

Since in the normal physiological condition, the production and removal of each component of the artery are balanced so that the vessel remains in its regular state and under a preferred homeostatic state, in the initial simulation, the material and geometric parameters are specified in such a way that AAA maintains its homeostatic mechanical state in the first 300 days. In the main simulation, the elastin damage function was applied to the abdominal aorta, leading to the degradation of elastin and a reduction in its quantity within the arterial wall, as described in Equation (1). Following the application of this degradation function, the simulation of AAA growth continued for an additional 3300 days. During this stage, elastin degradation causes an increase in the stress at the site of damage, according to

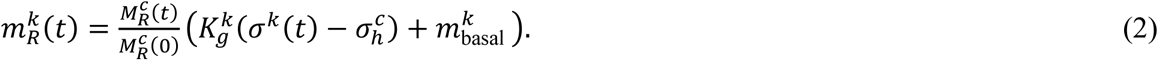

This leads to an increase in the production of collagen in the aortic wall, which has a compensatory role for the loss of elastin. In Equation (2), 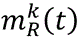 is the mass production rate of collagen fiber which proportional to a homeostatic stress 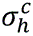; 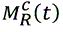 represents the total mass density of collagen fibers in the reference configuration, 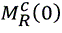 represents the mass density of collagen fiber in a healthy aorta at reference time 0; 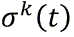 is stress on fiber family *k* at time *t*, and 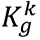 is a scalar parameter that controls the magnitude of stress-mediated mass production rate. The larger 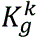 indicates that the blood vessel is capable of producing more collagen fibers to maintain mechanical stability under high vascular stress caused by elastin degradation. Therefore, 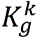 plays a crucial role in regulating the self-repair process and the evolution of blood vessels. Additionally, in this formula, 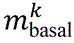 represents the basal production rate of collagen fibers [36–39].

The coefficients in Equations (1) and (2) play a significant role in the growth process and the final shape of the AAA. Given the importance of these coefficients and parameters in determining the final shape of the AAA, 80 trial simulations with different sets of coefficients were conducted to identify the plausible upper and lower limits of these coefficients, leading to the formation of more realistic and natural aneurysm shapes. After establishing the upper and lower bounds for each parameter, we used Latin Hypercube Sampling (LHS) to generate 200 parameter sets that uniformly span the design space. Each set populated the coefficients in Equations (1) and (2), and a full G&R simulation was then run for each, yielding 200 unique AAA growth trajectories and geometries. The simulations were conducted on a system with an 8-core processor, 3.8 GHz frequency, and 32 GB of memory. The computational duration of each G&R simulation varied between 7 and 11 hours, depending on the parameter values and the convergence behavior of the equations.

#### 2.1.4. Geometric Characteristics and Growth Rate of AAA

Following completion of the step of G&R simulation step, the growth rate, tortuosity, and maximum diameter of each AAA simulation were extracted using the Inscribed Maximal Diameter Curves (IMDCs) [40]. In this method, the AAA wall is considered a three-dimensional point cloud, and a sphere with the maximum inscribed diameter within the artery’s surface is defined. By moving the sphere from the beginning to the end of the artery, the central axis of the aneurysm is obtained, formed by the central points of the spheres. The geometrical parameters of the G&R simulation were recorded at 50-day intervals from the initial day to the 3600^th^ day, and the maximum diameter curve for each AAA was obtained at different time points.

The tortuosity of AAA is a geometrical parameter that is significant but uncorrelated with the volumetric variables (such as maximum diameter or volume) [41]. This parameter is defined as the ratio of the central axis curve length to the straight-line length between the start and end points of the aneurysm’s central axis. Another parameter that is particularly important in analyzing the growth pattern of AAAs is the growth rate, which can be calculated in both linear and exponential forms. To calculate these parameters, the maximum diameter of the aneurysm was first determined at each time point. Then, by using curve fitting methods, the linear and exponential growth rates for each curve were obtained.

The nonlinear growth rate was calculated using the following exponential function,

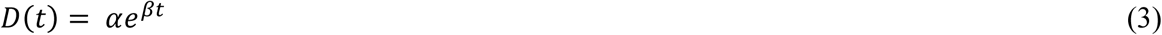

where *D*(*t*)is the diameter at time *t* (months); *ɑ* represents the initial maximum diameter, and *β* indicates the growth rate of the diameter. The other method for calculating the AAA growth rate is linear, which is computed as follows,

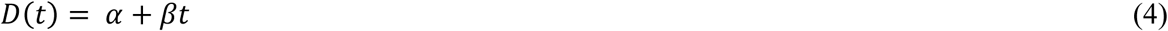

where *t* is measured in years, and *β* indicates the growth rate of the diameter. In these methods, *ɑ* and *β* are parameters that each have a specific value corresponding to the growth of a particular patient’s AAA [42,43], the time units are retained to remain consistent with the original formulations.

### 2.2. Generating Data Related to AAA Characteristics Using Surrogate Models

Because training the AAA growth prediction model based on machine learning methods requires a large dataset (tens of thousands samples) related to the aneurysm geometry and growth, and given the fact that performing finite element simulations of the AAA G&R for thousands of cases is computationally infeasible, the kriging statistical model [44] was used as a surrogate model to generate data related to the shape and growth of AAA. Kriging, also known as Gaussian process regression, provides the best unbiased linear prediction at unsampled locations. The results of G&R modeling were used as input data in the surrogate model.

Physics-based G&R simulations were performed for 200 parameter sets to generate AAA growth trajectories. To use the kriging method, the results from the G&R simulations must be divided into training and test datasets. The inputs of the kriging model included parameters related to the elastin degradation function in Equation (1) and the collagen production rate (Kg) in Equation (2). Based on these inputs, a kriging model was trained for each characteristic of aneurysm growth and geometry (growth rate, tortuosity, and maximum diameter) at different time intervals. Clinical CT imaging data from 25 patients were processed using the same feature-extraction pipeline, yielding 54 temporal sequences. Machine learning models were pre-trained using synthetic sequences and fine-tuned with clinical data to predict future maximum diameter and growth rate. Among the various kriging methods, the gradient-enhanced kriging with partial least squares (GEKPLS) method[45] was used due to its higher accuracy and speed compared to standard kriging.

### 2.3. Machine Learning Models to Predict AAA Growth

In order to predict the growth of AAAs, four different ML models, including Deep Belief Network (DBN), Recurrent Neural Network (RNN), Long Short-Term Memory (LSTM) network, and Gated Recurrent Unit (GRU) network, were utilized in this study. The performance of these models in predicting the growth and geometry characteristics of AAAs was compared. In the previous section (2.2), the kriging method was used as a technique for reproducing a large amount of in-silico data, including maximum diameter, tortuosity, and growth rate, across 6-time points of aneurysm growth (from 2100^th^ day to 3600^th^ day, sampled at 300-day intervals to align with the spacing of the clinical follow-up data). Subsequently, to align the temporal structure of the in-silico trajectories with that of clinical imaging data, which typically consist of approximately four longitudinal scans, we re-formatted each 6-time-point simulation into three overlapping 4-time-point sequences. In this study, a “sequence” refers to a set of consecutive geometric measurements sampled at fixed temporal intervals (300 days). This ensured that the model inputs mirrored the length and spacing of real patient follow-up data. After constructing these aligned subsequences, 30% were allocated to validation and testing, and the remainder were used for training.

These machine learning models operate on time-series representations of aneurysm geometry extracted from either G&R simulations or clinical imaging data. At each observation time point, three scalar geometric descriptors are computed: maximum diameter, tortuosity, and diameter growth rate. These quantities are obtained using the IMDC-based post-processing pipeline described in Section 2.1.4. For each simulated or clinical aneurysm trajectory, measurements at six time points are reorganized into overlapping sequences of four consecutive time points. The prediction target corresponds to the next time step. Thus, the machine learning models learn a temporal mapping from prior aneurysm geometry evolution to future aneurysm diameter and growth rate.

The two main innovations in the ML section of this study are as follows:

1. Generating a large in silico dataset using physics-based computational models and approximation algorithms to address the scarcity of real patient data.
2. Introducing a deep learning-based two-step training method, which combines real patient data and synthetic in silico data to improve prediction accuracy.

#### 2.3.1. Deep Belief Network

A two-stage learning scheme is used to train the deep belief network [46]. In summary, the network was initially pre-trained with in silico data using the Contrastive Divergence (CD) method in an unsupervised manner. Then, the network was fine-tuned with labeled patient data as standard neural network.

During the pre-training phase of the DBN, the initial 3 sequences of maximum diameter, tortuosity, and growth rate were used as visible units in the first layer of a restricted Boltzmann machine (RBM). It should be noted that Gaussian RBMs were used in all layers. Upon completion of the pre-training step, the DBN was converted into a standard neural network with an additional linear layer consisting of two output nodes for supervised training. In this model, the values of the two output nodes were used to predict the final sequence values of the maximum diameter and growth rate of the aneurysm.

#### 2.3.2. Recurrent Neural Network

RNN is a type of artificial neural network used for predicting sequential data. In RNNs, there is a feedback layer where the network’s output, along with the next input, is fed back into the network. This feedback mechanism allows the RNNs to retain information from previous inputs through internal memory, allowing them to learn and process sequences of inputs [47].

In addition to the simple RNN, there are other types, such as LSTM networks and GRU networks [48,49]. These variations modify the blocks of the network to address issues like vanishing or exploding gradients. These networks are typically used for predicting long sequences, but they can also perform better than simple RNNs in some cases with shorter sequences.

Given the sequential nature of the data, including maximum diameter, tortuosity, and growth rate at various time intervals, recurrent models can provide more accurate predictions of maximum diameter and growth rate compared to standard neural networks by understanding the temporal relationships between the data. For training the models, 70% of the data was used in the form of matrices (m×n, where m is the number of sequences and n is the number of input features — maximum diameter, tortuosity, and growth rate), while the remaining data was reserved for testing and validation. To enable comparison between different types of RNN models, the number of layers and nodes in each of the three RNN models was kept the same. In the process of training, three primary time sequences of data (maximum diameter, tortuosity, and growth rate) were used as model input, while values of maximum diameter and growth rate in the last time sequence were used as output.

#### 2.3.3. Fine-Tuning with Patient Data

In machine learning, fine-tuning is an approach for transfer learning where the weights of a pre-trained model are adjusted using new data. Fine-tuning can be performed on the entire neural network or just on a subset of its layers, and it is crucial for improving accuracy and enhancing model predictions on real-world data. In this study, fine-tuning was performed using medical imaging data from 25 patients acquired at multiple time points. The study was determined to be non-human subjects research by the Institutional Review Board at Michigan State University, as the dataset was retrospectively collected and deidentified. To convert the medical imaging data into numerical aneurysm growth characteristics, the IMDCs method was employed to extract the maximum diameter and centerline geometry. Tortuosity was computed from the centerline, and growth rate was estimated from sequential diameter measurements using linear regression over fixed 300-day intervals. Growth rate is computed from prior diameter evolution and is treated as a derived geometric descriptor summarizing recent expansion trends, allowing the model to incorporate both instantaneous geometry and temporal change information. Given that the time intervals between medical images for each patient are variable, interpolation was used to calculate the characteristics of maximum diameter, tortuosity, and growth rate of the AAA at a fixed time interval (300 days). After interpolation, 54 sequence sets (each sequence consists of data in four successive time intervals) with a fixed interval of 300 days for each of the characteristic maximum diameter, tortuosity, and growth rate, were obtained. Out of these, 8 sequences were set aside as test data, and 46 sequences were used for the fine-tuning process. To evaluate the influence of temporal input length on prediction performance, additional experiments were conducted using sequences composed of one, two, and three prior time points. In each case, the same dataset, model architecture, and training procedure were used, while only the number of input time steps was varied. The patient data are listed in **Table 1**. All four models used (the DBN and the three RNN models) were fine-tuned with these patient data. **Figure 2** shows the overall schematic of the steps carried out in this study.

**Figure 2.**
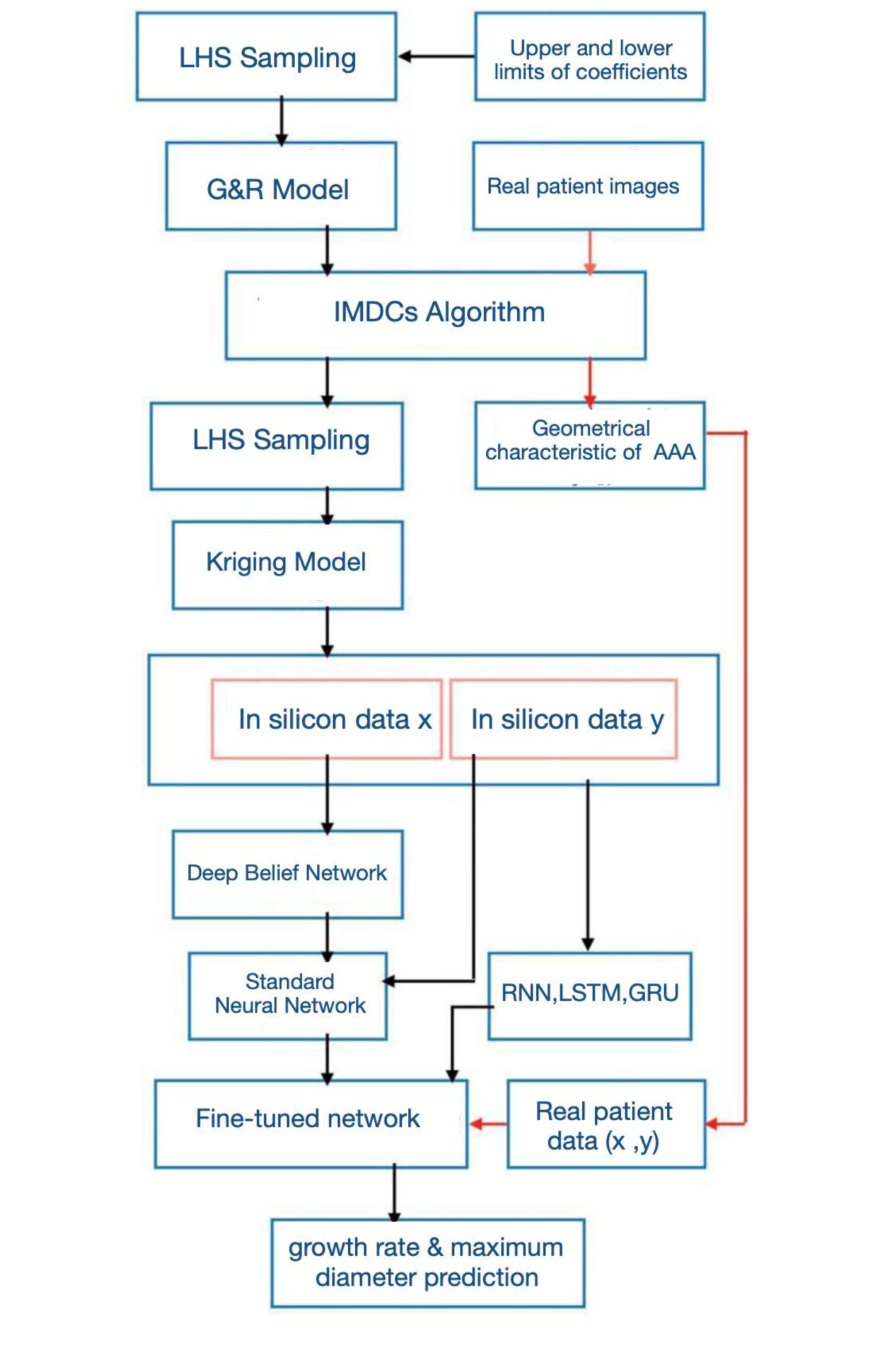
Diagram of the modeling carried out in this study.

**Table 1.** Data of patients.

| Patient | # Scan | Gender | Time of scans (days) |
| --- | --- | --- | --- |
| 1 | 4 | M | [0, 208, 733, 943] |
| 2 | 4 | F | [0, 404, 708, 1357] |
| 3 | 5 | M | [0, 182, 361, 538, 728] |
| 4 | 5 | M | [0, 347, 702, 1054, 1223] |
| 5 | 5 | M | [0, 374, 1074, 1438, 2136] |
| 6 | 5 | M | [0, 386, 757, 1121, 1290] |
| 7 | 5 | M | [0, 227, 674, 1049, 1403] |
| 8 | 3 | M | [0, 97, 564] |
| 9 | 4 | M | [0, 522, 922, 1344] |
| 10 | 4 | M | [0, 399, 774, 1152] |
| 11 | 3 | M | [0, 523, 873] |
| 12 | 4 | M | [0, 543, 691, 874] |
| 13 | 3 | M | [0, 349, 714] |
| 14 | 5 | M | [0, 183, 366, 534, 709] |
| 15 | 3 | M | [0, 189, 826] |
| 16 | 5 | M | [0, 246, 421, 587, 783] |
| 17 | 4 | F | [0, 613, 1048, 1515] |
| 18 | 4 | M | [0, 309, 1976, 2310] |
| 19 | 4 | M | [0, 729, 922, 2359] |
| 20 | 3 | M | [0, 440, 990] |
| 21 | 5 | M | [0, 343, 610, 1480, 2232] |
| 22 | 4 | F | [0, 401, 988, 1504] |
| 23 | 4 | F | [0, 167, 543, 920] |
| 24 | 3 | M | [0, 442, 880] |
| 25 | 4 | M | [0, 243, 730, 1502] |

## 3. Results

The results of this study are structured into three parts: (1) G&R simulations of AAAs and calculating maximum diameter, tortuosity, and growth rate values at different time intervals for 200 different parameter sets, including elastin degradation coefficients and collagen production rate. (2) Using the results of the G&R simulations step as input data for the statistical model (kriging) and utilizing this model to generate the required sequences of maximum diameter, tortuosity, and growth rate for training machine learning models. (3) Implementing a two-stage learning scheme consists of pre-training and fine-tuning. In the pre-training part, four different models (DBN, RNN, LSTM, and GRU) were utilized, and the performance of these models in predicting the growth characteristics and geometry of AAAs was evaluated.

### 3.1. G&R Simulations Results

The G&R simulation results were transferred to Paraview software (Kitware Inc., Clifton Park, NY, USA) to evaluate stress, mass density rate of collagen, and geometrical characteristics of AAAs. **Figure 3** shows the shape evolution and the von Mises stress distribution contour of AAA for one of the simulations at different time intervals (1200^th^ to 3600^th^ day). Due to increased stress in regions where the elastin degradation function is applied, arterial dilation is observed in these areas. According to Eq. 2 in these areas, the mass density of collagen increases in order to reduce the effect of elastin degradation. **Figure 3** presents the contours of collagen mass density rates.

**Figure 3.**
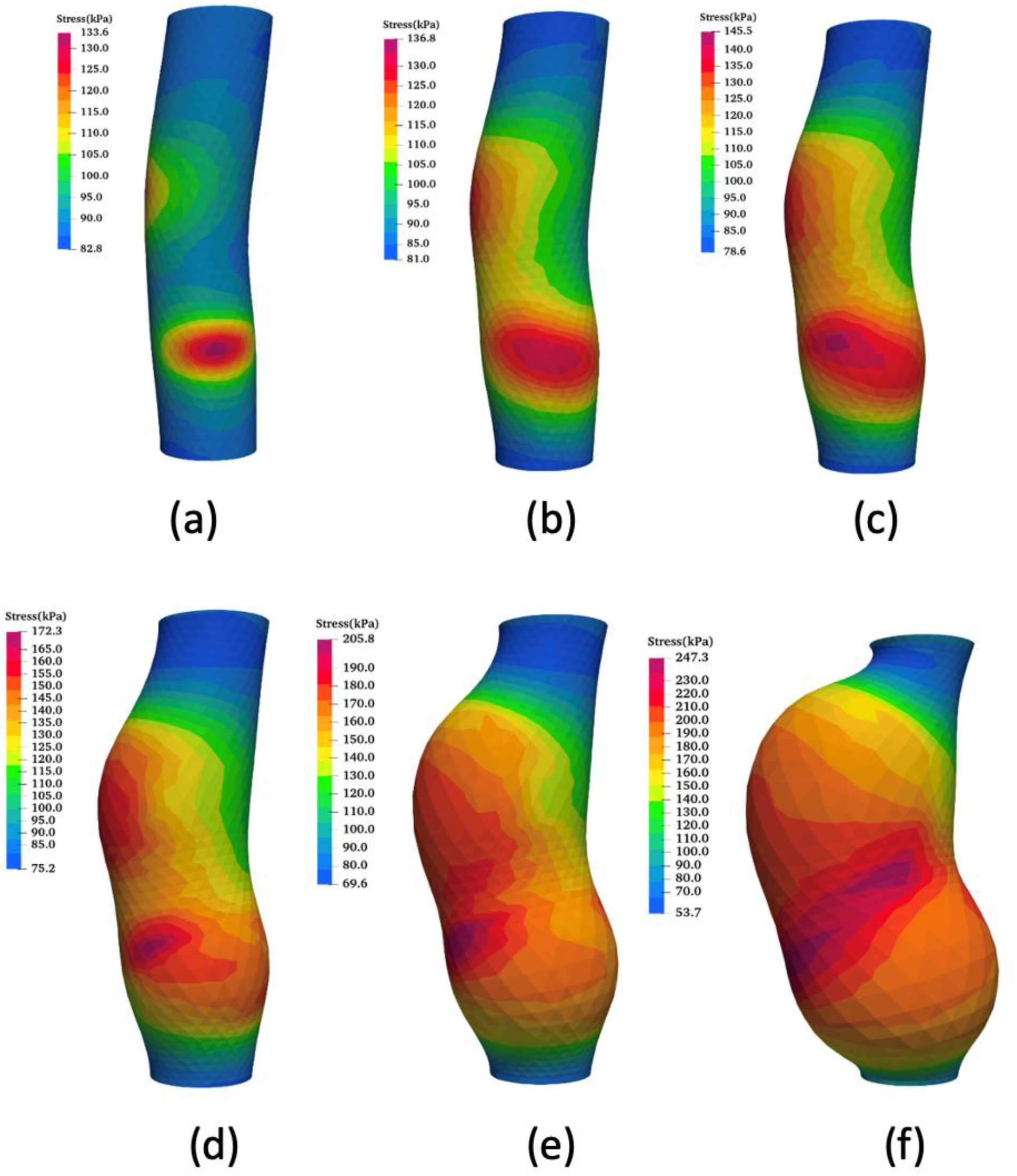
Distribution of von Mises stress (kPa) for AAA with a specific set of parameters at different time intervals: (a) 300 days, (b) 1200 days, (c) 1800 days, (d) 2400 days, (e) 3000 days, and (f) 3600 days.

Collagen is one of the main components that withstands the mechanical load in the artery wall. As shown in **Figures 3** and **4**, an increase in stress will activate the compensation mechanism of collagen so that the artery can produce more collagen to bear mechanical forces. However, the relationship between the amount of stress and the rate of collagen production is not linear, and many other factors are effective in this process.

**Figure 4.**
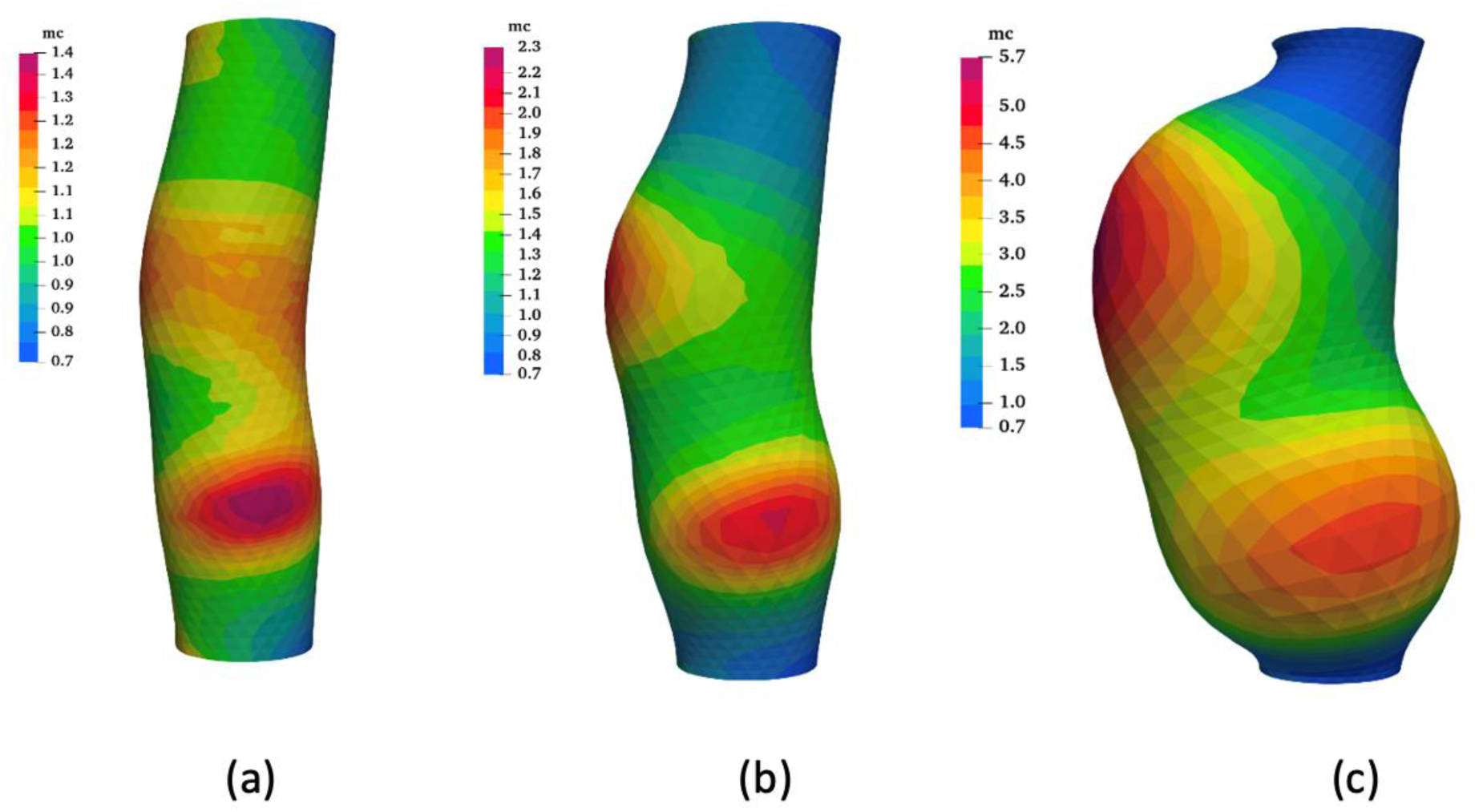
Mass density of collagen distribution (kg/m²) at different time intervals: (a) 1200 days, (b) 2400 days, and (c) 3600 days.

In **Figure 5**, twelve different morphologies of simulated AAAs are shown. As it can be seen, the simulations with different parameter sets have been able to create an acceptable variety in the growth rate and geometry of the aneurysm while producing realistic shapes that have a high agreement with the medical images.

**Figure 5.**
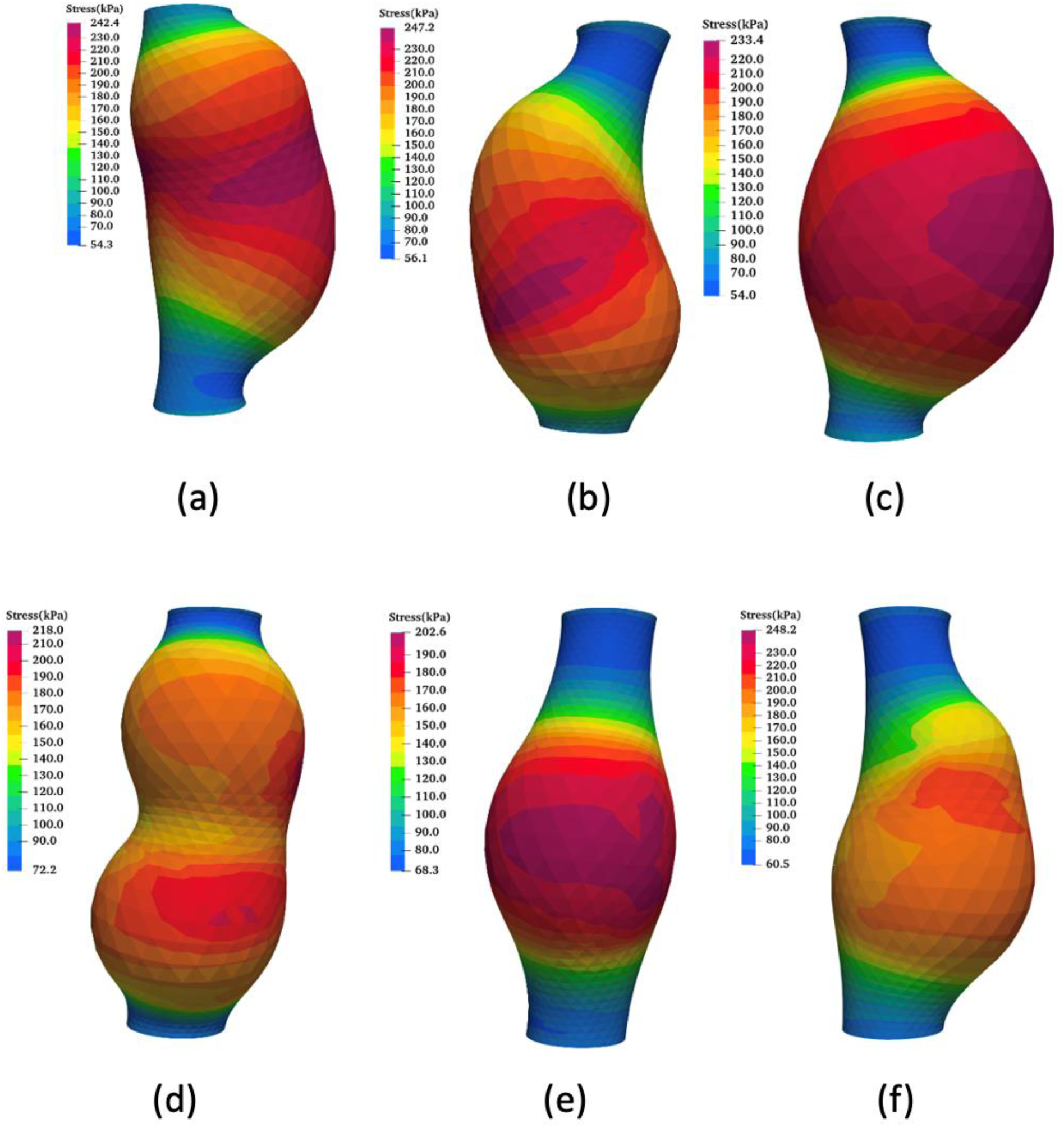
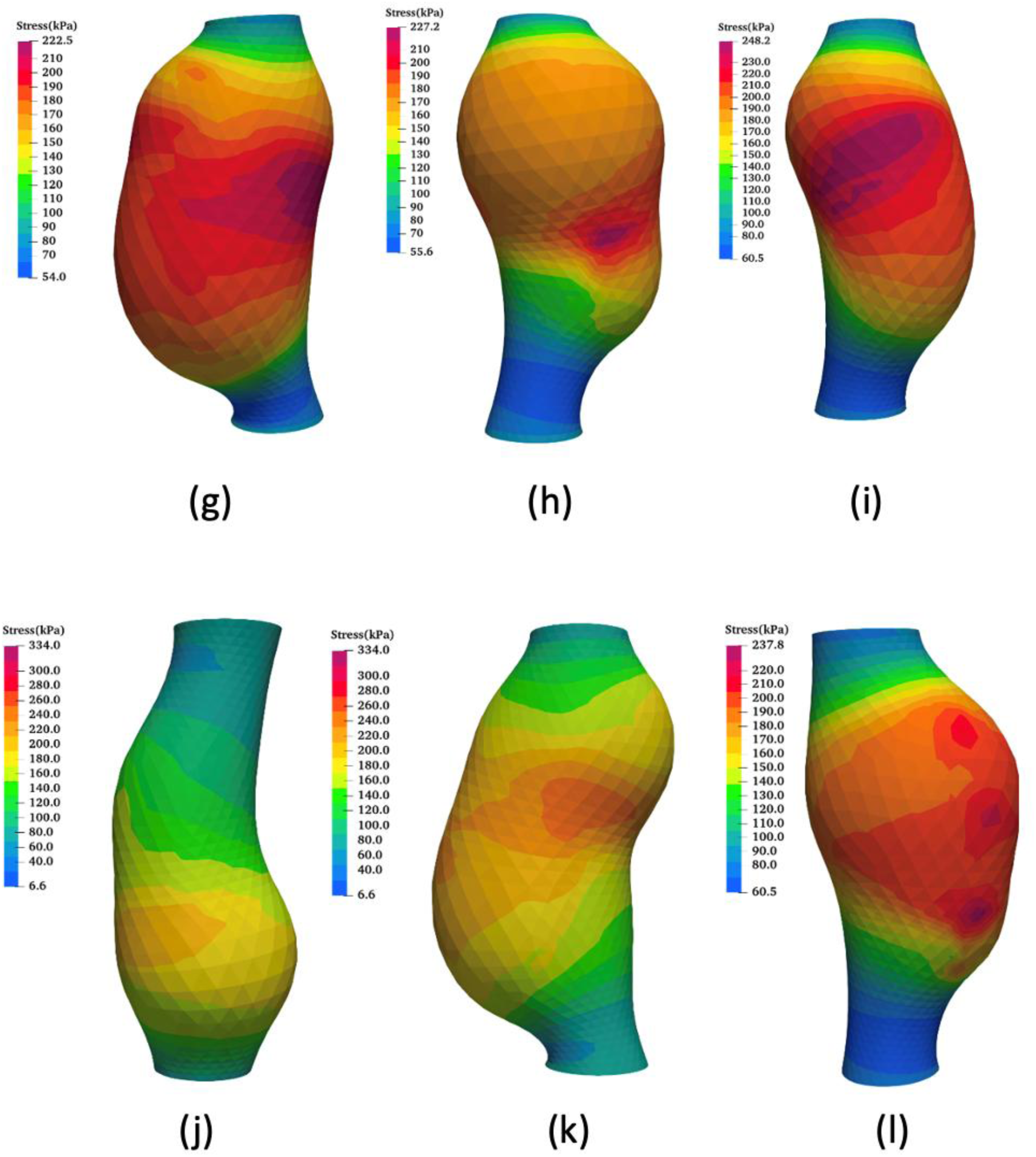
Twelve different morphologies of AAAs.

In the post-processing step, the maximum diameter, tortuosity, and growth rate were extracted from the outputs of the 3D simulations. The IMDC technique was used to obtain the maximum diameter of each aneurysm at different time intervals. In **Figure 6**, the diameter changes along the center axis for two different aneurysms are depicted.

**Figure 6.**
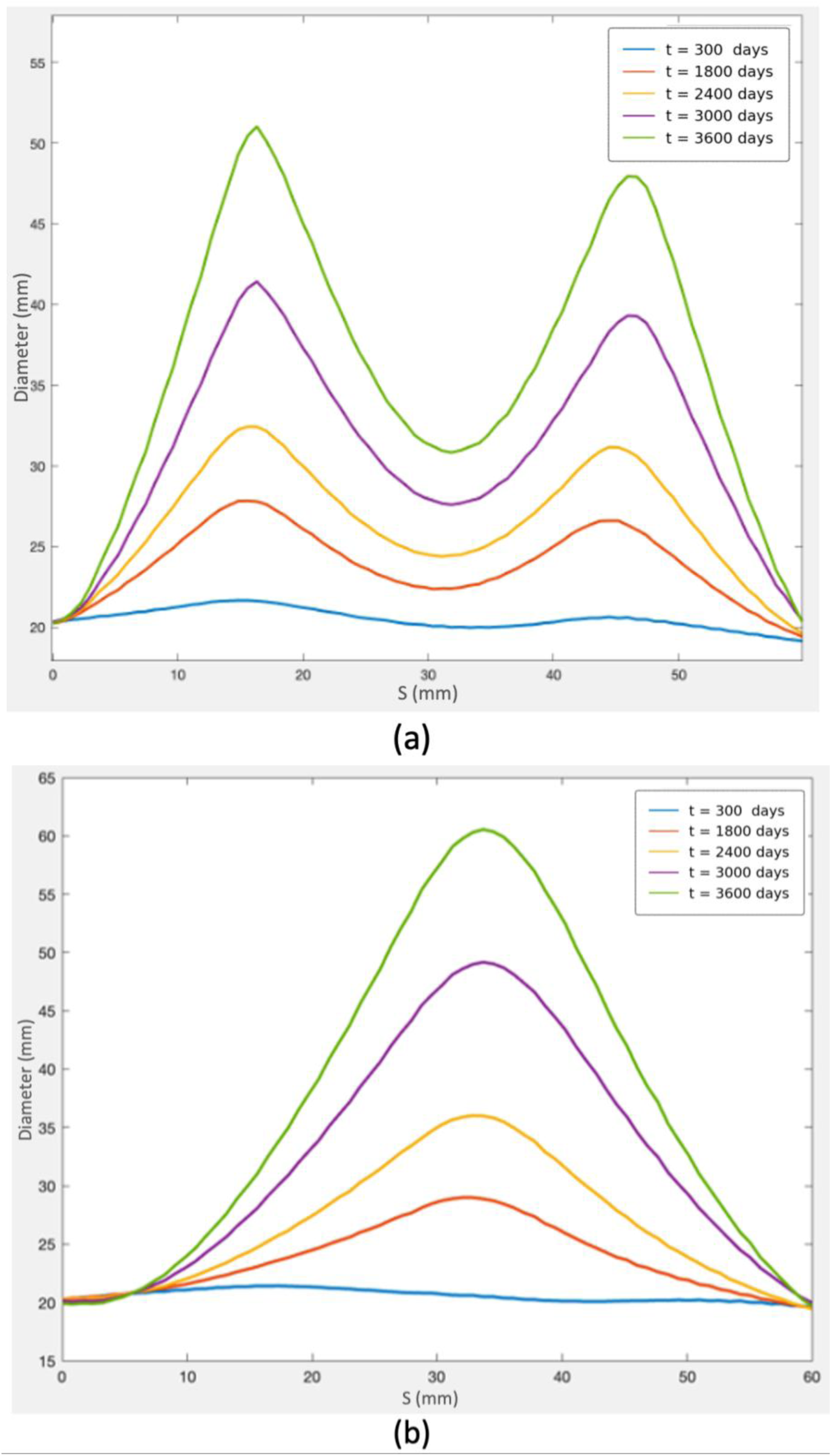
Maximum diameter profiles at different times along the center axis of the artery.

In the next step, these data were applied to fit growth rate curves, as shown in **Figure 7**, which presents an example of fitting both linear and exponential growth rate models. It is worth noting that the exponential model generally achieves better accuracy for long-term intervals, while the linear model is more accurate in short-term intervals.

**Figure 7.**
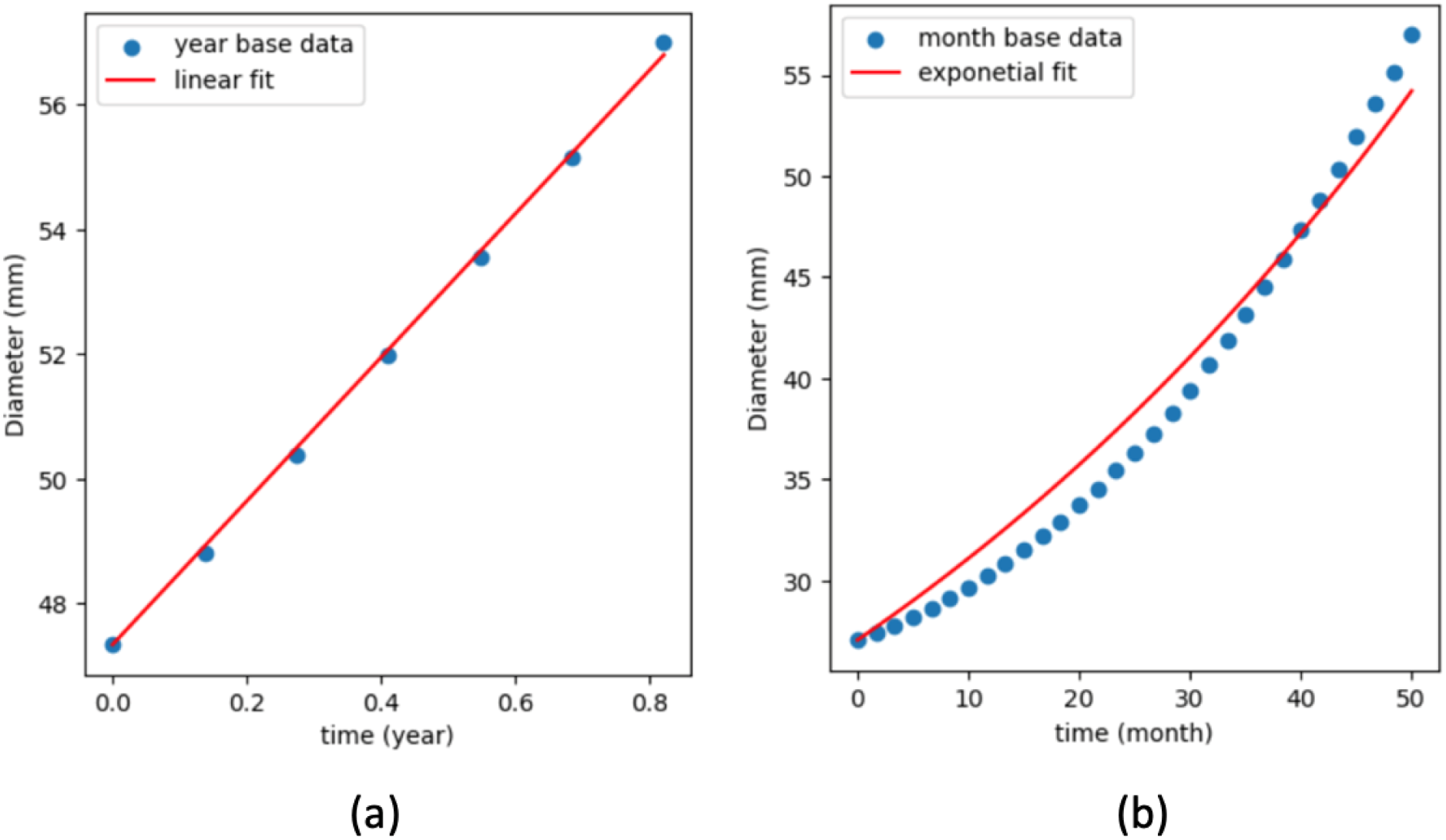
Growth rate: (a) Linear and (b) Exponential.

In **Figure 7(a)**, the linear growth rate curve fitting with simulation data is shown for maximum diameter data within a 300-day period (from day 3300 to day 3600). As observed, the data exhibit linear behavior within this time frame, making the linear growth rate a more suitable choice. In **Figure 7(b)**, the exponential growth rate curve fitting is displayed for maximum diameter data within a 1500-day period (from day 2100 to day 3600). According to the goal of this study, using shorter time intervals for predicting growth rate and maximum diameter in subsequent intervals, the linear growth rate was employed in the following steps.

In order to differentiate between AAAs, tortuosity as another criteria (in addition to maximum diameter and growth rate) was evaluated, utilizing the IMDC method. The resulting tortuosity values ranged from 1.01 to 1.20, which is consistent with ranges reported in previous clinical and computational studies [24,50].

In the next step, the collagen production rate coefficient, *Kg*, was investigated as a parameter that controls collagen production based on stress values, and its relation with the growth and shape changes of AAA was evaluated. For this purpose, different values for *Kg* were considered, assuming the other parameters to be constant, and the maximum diameter of the aneurysm was calculated at each time interval. **Figure 8** shows the maximum diameter of the aneurysm at different time intervals. Lower values of *Kg* cause faster and greater enlargement of the maximum diameter. Since the reduction of *Kg* values reduces the effect of the collagen compensatory mechanism, and due to the destruction of elastin in the artery wall and insufficient production of collagen, the artery wall expands to larger values. These results are consistent with previous studies [41,43,51].

**Figure 8.**
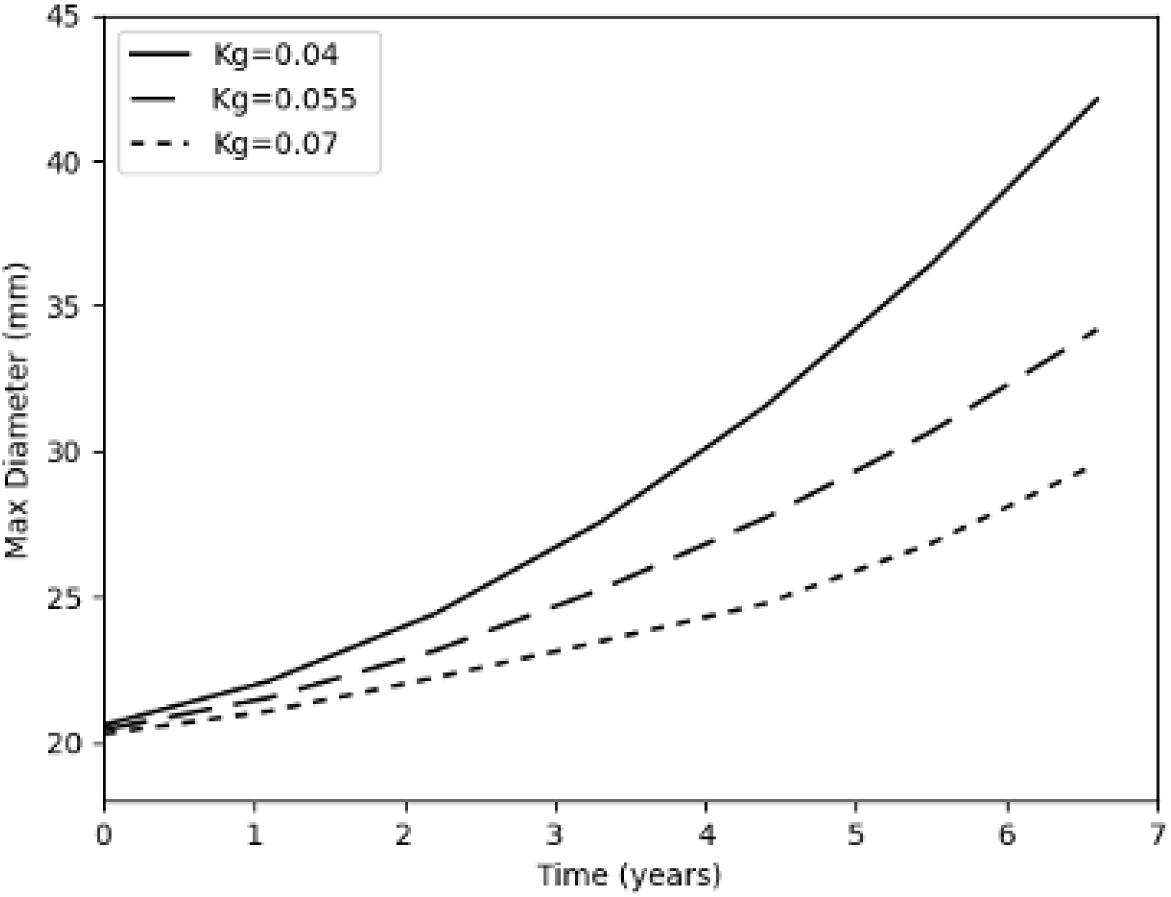
The maximum diameter of AAAs at different Kg values plotted against time.

It should be noted that the previous studies showed that the variation in *Kg* values is due to patient-specific genetic factors or individual lifestyle differences. Therefore, it may be possible to explain the change in collagen values observed among different AAAs for this reason.

### 3.2. Kriging Model Results

The output of 3D G&R modeling, including the maximum diameter, growth rate, and tortuosity for 200 different simulations, was used as input data for the GEKPLS model to generate the required training data for the machine learning step. In **Figures 9-11**, the output curves at day 3600^th^ from this model, along with the 95% confidence intervals for each characteristic (maximum diameter, growth rate, and tortuosity) are presented in response to a change in each parameter defined in Eq. 1, while the other input parameters remain constant. As illustrated, each parameter affects the aneurysm’s shape and size differently. For instance, the parameter *Kg* significantly influences the aneurysm’s maximum diameter, with higher values leading to a noticeable decrease in maximum diameter. On the other hand, the parameters related to the damage location along the central axis (μ_*s*_ and μ_*θ*_) have a lesser effect on the maximum diameter but considerably influence the aneurysm’s final shape, resulting in aneurysms with distinct shapes based on parameter variations.

**Figure 9.**
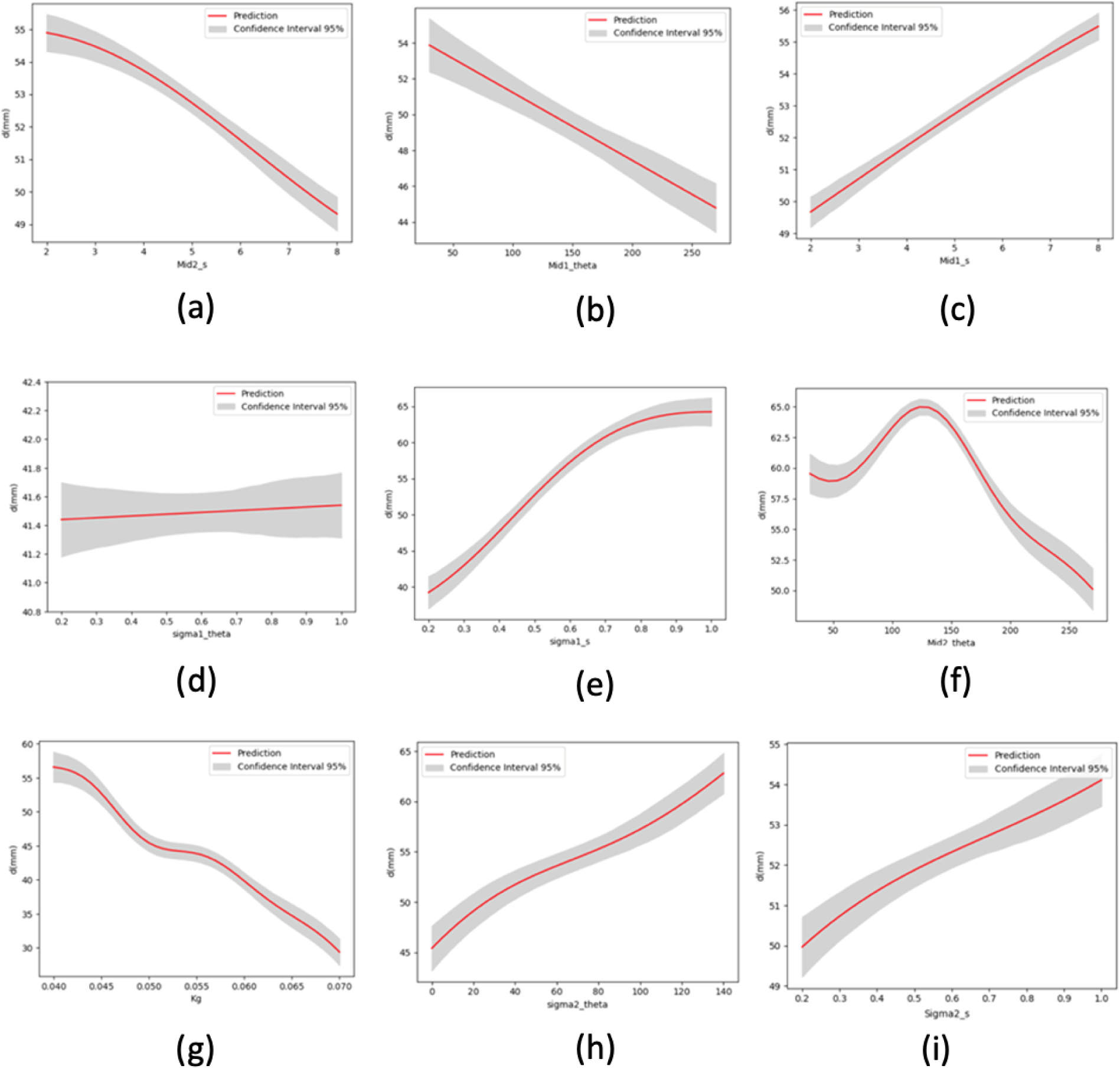
Diameter curves using the Kriging model based on variations in each input parameter while keeping the other parameters constant.

**Figure 10.**
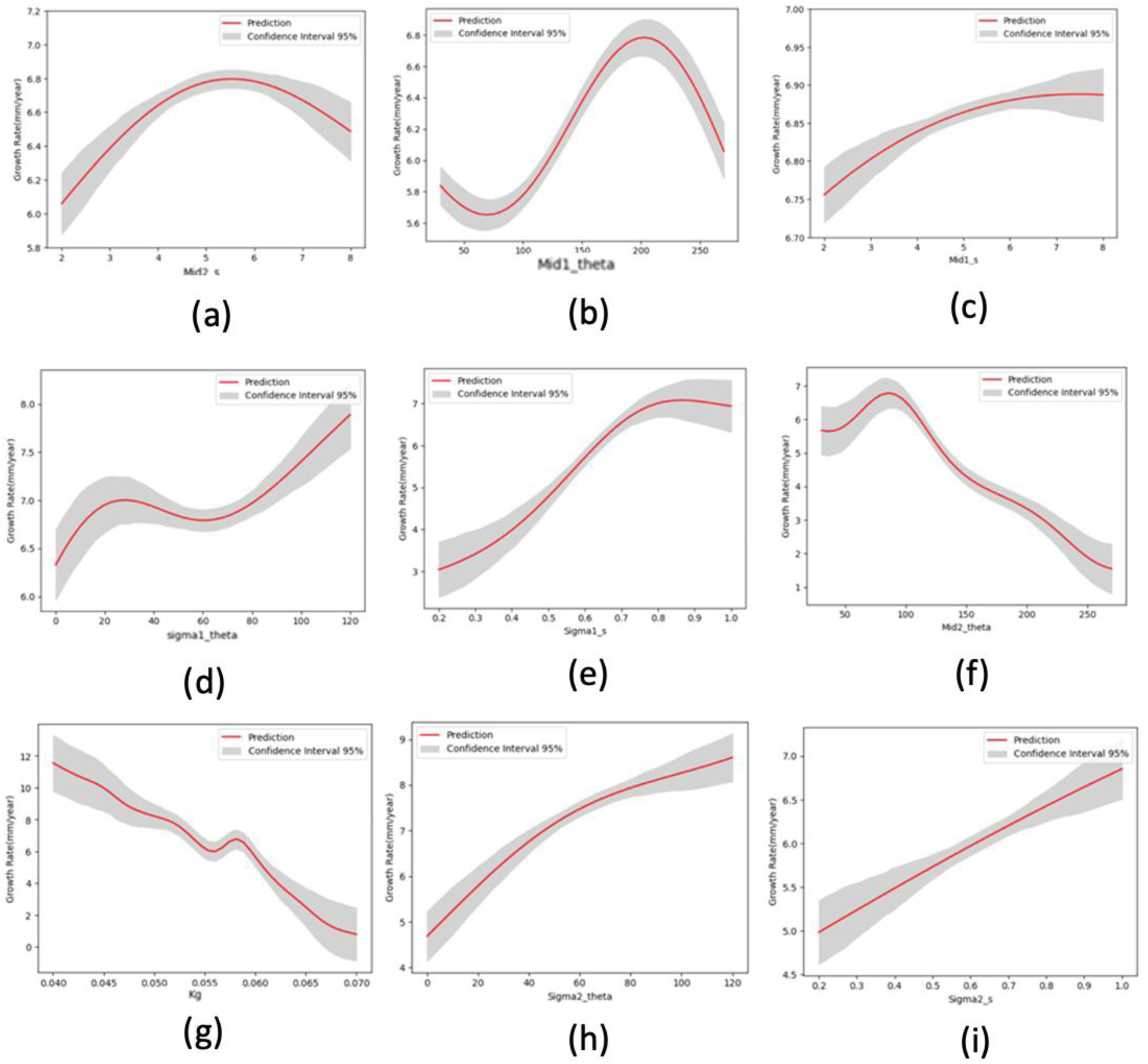
Growth rate using the Kriging model based on variations in each input parameter while keeping the other parameters constant.

**Figure 11.**
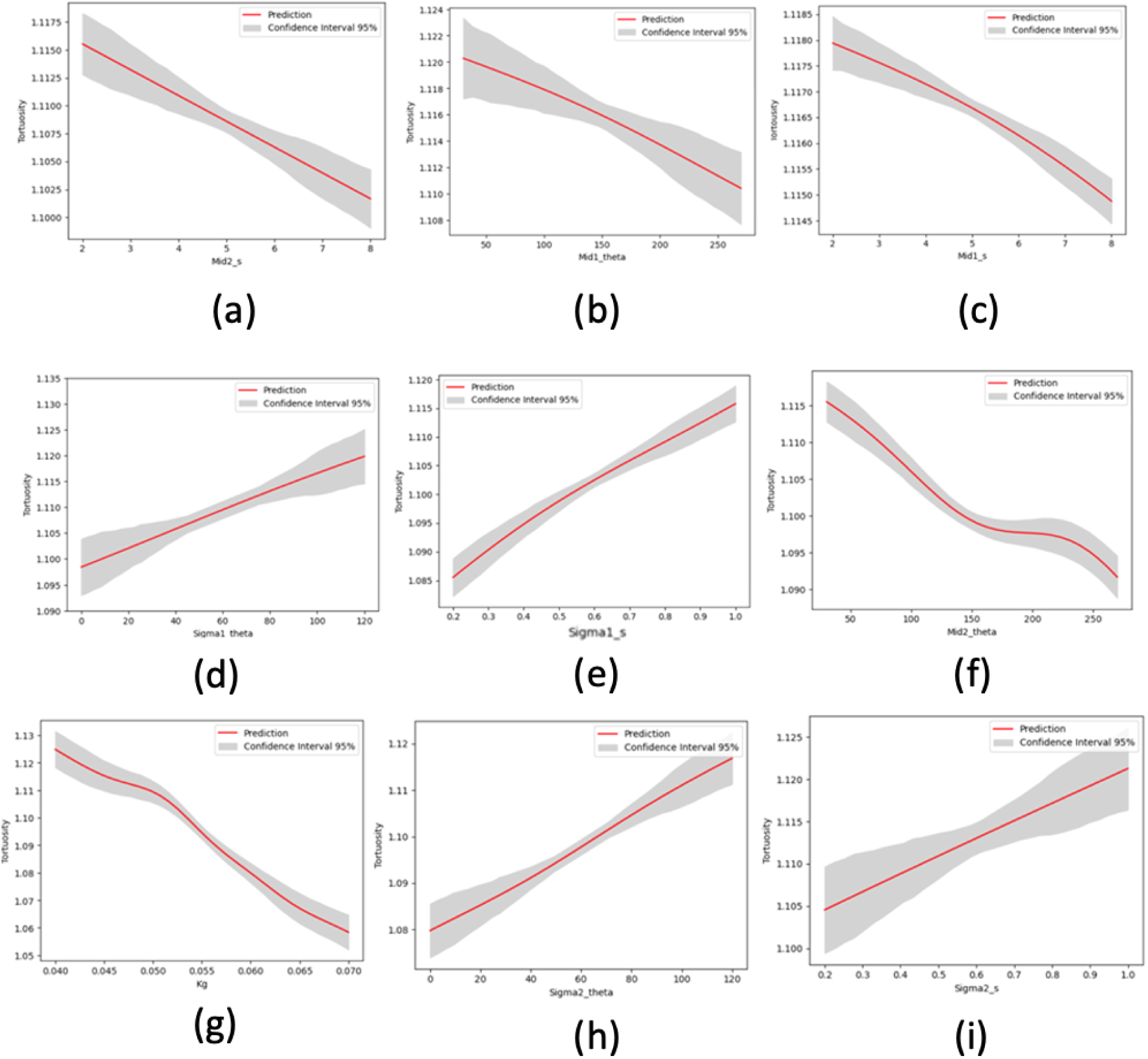
Tortuosity using the Kriging model based on variations in each input parameter while keeping the other parameters constant.

To evaluate the predictive accuracy of the GEKPLS surrogate model, 20% of the geometric and growth data from the G&R simulations, which were not included in the training phase of the GEKPLS model, were utilized to calculate the root mean square error (RMSE) values. RMSE values were computed for key features of AAA progression: maximum diameter, growth rate, and tortuosity. These metrics quantify prediction errors by measuring the discrepancy between actual values from G&R simulations and the predicted values generated by the GEKPLS model.

In terms of maximum diameter, the model achieved an RMSE of 3.6, which is small relative to the typical AAA diameter range (∼30–60 mm). For growth rate, the RMSE was 1.5, representing a modest error compared with commonly reported annual growth rates. In contrast, the RMSE for tortuosity was 0.011; although numerically smaller, this value is appropriate given that tortuosity itself varies over a narrow range (approximately 1.01–1.20), making the relative error similarly low. These results demonstrate that the GEKPLS surrogate model is capable of replicating the key characteristics predicted by the computationally intensive G&R simulations within an acceptable margin of error (**Table 2**).

**Table 2.** RMSE of the Kriging model.

| Parameter | RMSE |
| --- | --- |
| Diameter (mm) | 3.6 |
| Growth rate (mm/year) | 1.5 |
| Tortuosity (-) | 0.011 |

### 3.3. Results of Machine Learning Models

At this stage, machine learning models were used to predict the maximum diameter, tortuosity, and growth rate of AAA. As mentioned before, to solve the limited clinical data problem, the output dataset of the G&R simulations was used as the input of the kriging method as an approximate method to reproduce a large amount of in silico data. Here, a two-stage learning scheme was used to train DBN, RNN, LSTM, and GRU networks.

Firstly, the performance of each model was evaluated on the in silico data. **Figure 12** shows the predicted maximum diameter versus the in silico data. According to this figure, the models have acceptable accuracy in predicting the maximum diameter. **Table 3** shows the R^²^ score and root mean square error for each of the models.

**Figure 12.**
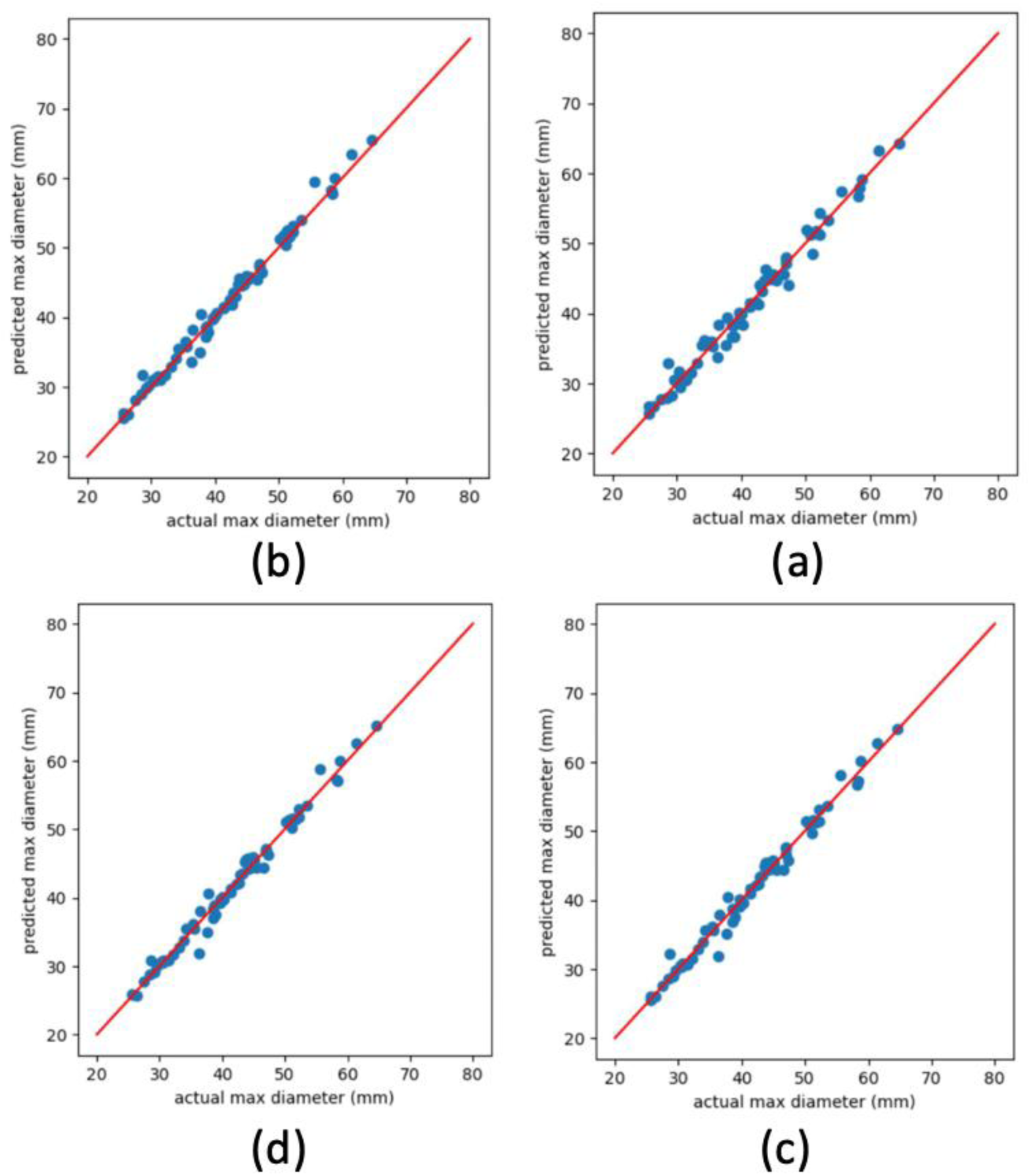
Predicted maximum diameter versus actual maximum diameter (in silico) by models a: DBN, b: RNN, c: LSTM, d: GRU.

**Table 3.** R² score and RMSE for different models in predicting maximum diameter (in silico).

|  | GPU | LSTM | RNN | DBN |
| --- | --- | --- | --- | --- |
| RMSE | 1.68 | 1.61 | 1.67 | 1.93 |
| $R^2$ score | 0.96 | 0.97 | 0.96 | 0.91 |

Given the limited number of patient dataset (*n*=54), the k-fold method was used to train and evaluate the model’s performance. In this method, all data are divided into *k* groups; in each iteration, *k*−1 groups are used to train the model, and one group is used to test the model’s performance. This process is repeated *k* times. It is worth noting that in this approach, no data used for training is utilized for performance testing, and in each iteration, the training and test data are completely separated. In **Figure 13**, the prediction graph of each model for the maximum diameter of AAA in comparison with the actual maximum diameter (patient data) is shown. In the RNN, LSTM, and GRU models, three previous time points (*t-3*, *t-2*, and *t-1)* sequences were used to predict the next sequence(*t*). **Figure 13** compares actual versus predicted maximum diameters (mm) for four machine learning models: RNN, DBN, LSTM, and GRU. The RNN and LSTM models demonstrate superior predictive accuracy, with data points closely clustering along the diagonal line, indicating minimal deviation and strong correlation between predictions and actual values across a range of diameters. In contrast, the DBN and GRU models exhibit greater variability, especially for larger diameters, with noticeable deviations from the diagonal line, suggesting reduced predictive reliability. Overall, the RNN and LSTM outperform the DBN and GRU.

**Figure 13.**
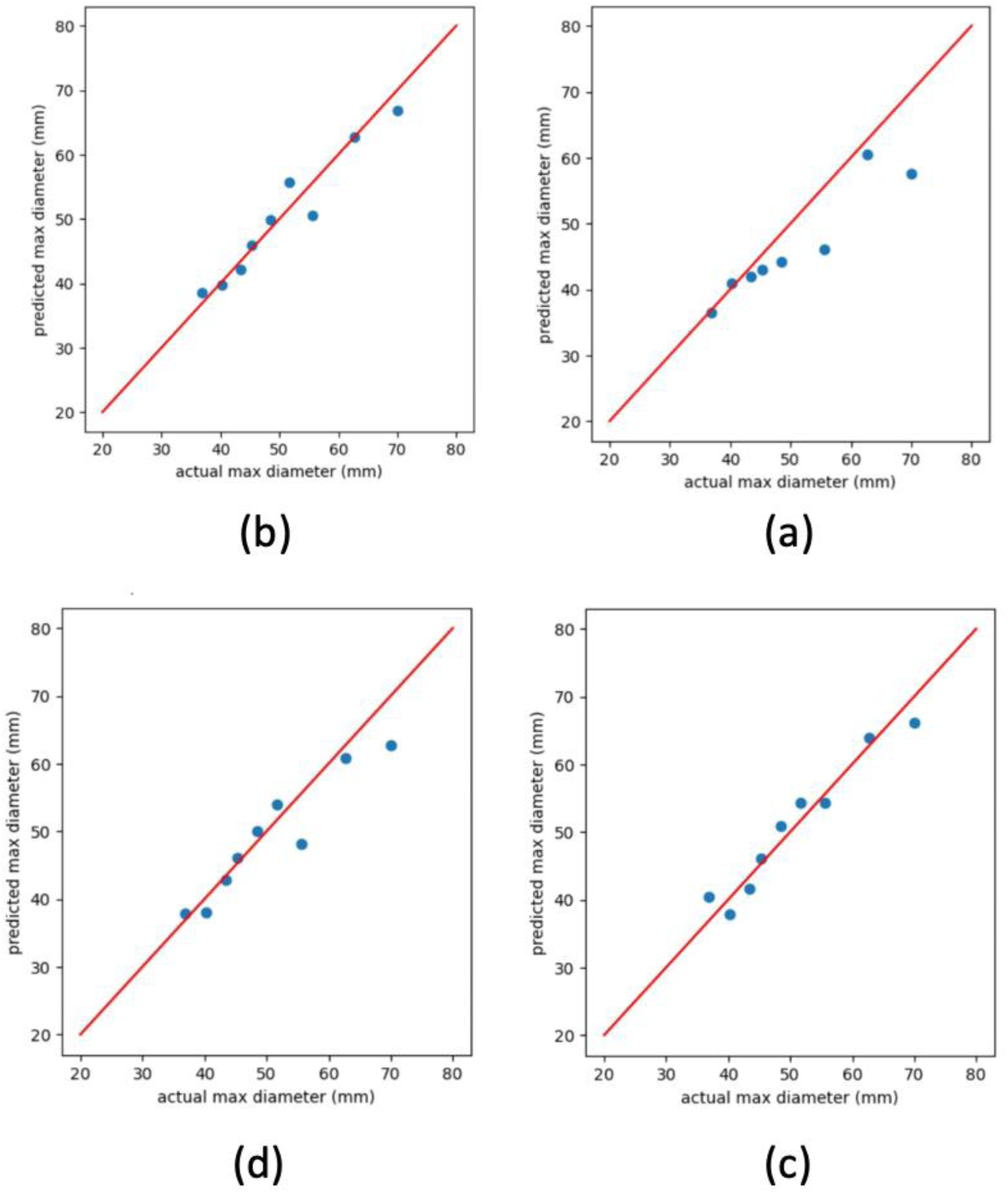
Predicted maximum diameter versus actual maximum diameter (selected patient dataset) by models a: DBN, b: RNN, c: LSTM, d: GRU.

**Table 4** evaluates the performance of four machine learning models (GRU, LSTM, RNN, and DBN) in predicting maximum aneurysm diameter using patient data. The LSTM model demonstrated the best predictive accuracy with the lowest RMSE (1.84 mm) and the highest R² score (0.92), indicating minimal error and strong correlation with actual values. The RNN performed comparably well, with an RMSE of 1.91 mm and an R² score of 0.90, followed by the GRU model, which showed slightly higher prediction error with an RMSE of 1.93 mm and an R² score of 0.89. In contrast, the DBN model had the weakest performance, with an RMSE of 2.47 mm and an R² score of 0.82, reflecting greater predictive error and weaker correlation.

**Table 4.** R² score and RMSE for different models in predicting maximum diameter (patient data).

|  | GPU | LSTM | RNN | DBN |
| --- | --- | --- | --- | --- |
| RMSE | 1.93 | 1.84 | 1.91 | 2.47 |
| $R^2$ score | 0.89 | 0.92 | 0.90 | 0.82 |

**Table 5** illustrates the R² scores of three recurrent machine learning models (GRU, LSTM, and RNN) in predicting maximum aneurysm diameter across different sequence lengths (Three Sequence, Two Sequence, and One Sequence). The results demonstrate that prediction performance improves with longer sequence input data, as indicated by increasing R² scores across all models. For three sequence inputs, the LSTM model achieved the highest R² score of 0.92, showcasing its superior ability to learn temporal dependencies over extended sequences, followed by the RNN with 0.90 and GRU with 0.89. As the sequence length decreased, model performance gradually declined; for two sequence inputs, the R² scores dropped to 0.86 for both LSTM and RNN, and 0.84 for GRU. With only one sequence, the models exhibited the lowest predictive accuracy, with LSTM scoring 0.79, followed by RNN at 0.77 and GRU at 0.73. These findings highlight the importance of incorporating longer temporal sequences to enhance predictor reliability in recurrent models, with LSTM consistently outperforming GRU and RNN across all sequence lengths.

**Table 5.** R² score of recurrent models in predicting maximum diameter for different sequence lengths.

| $R^2$ score | GRU | LSTM | RNN |
| --- | --- | --- | --- |
| Three sequence | 0.89 | 0.92 | 0.90 |
| Two sequence | 0.84 | 0.86 | 0.86 |
| One sequence | 0.73 | 0.79 | 0.77 |

According to **Figure 14**, the LSTM model demonstrated the best performance, as the predicted growth rates were most tightly clustered along the diagonal, highlighting its ability to predict growth trends with minimal error and high correlation with actual values. The RNN model (**Fig. 14b**) also exhibited strong predictive capability, with data points showing moderate clustering near the diagonal and low variance, indicating reliable predictions across the dataset. In contrast, the GRU model (**Fig. 14d**) and DBN model (**Fig. 14a**) showed weaker predictive performance relative to LSTM and RNN. The data points for GRU displayed larger dispersion, deviating significantly from the ideal diagonal in several cases, reflecting lower precision and increased prediction error. The DBN model displayed wider deviations from the diagonal line, suggesting substantial inaccuracies in growth rate prediction.

**Figure 14.**
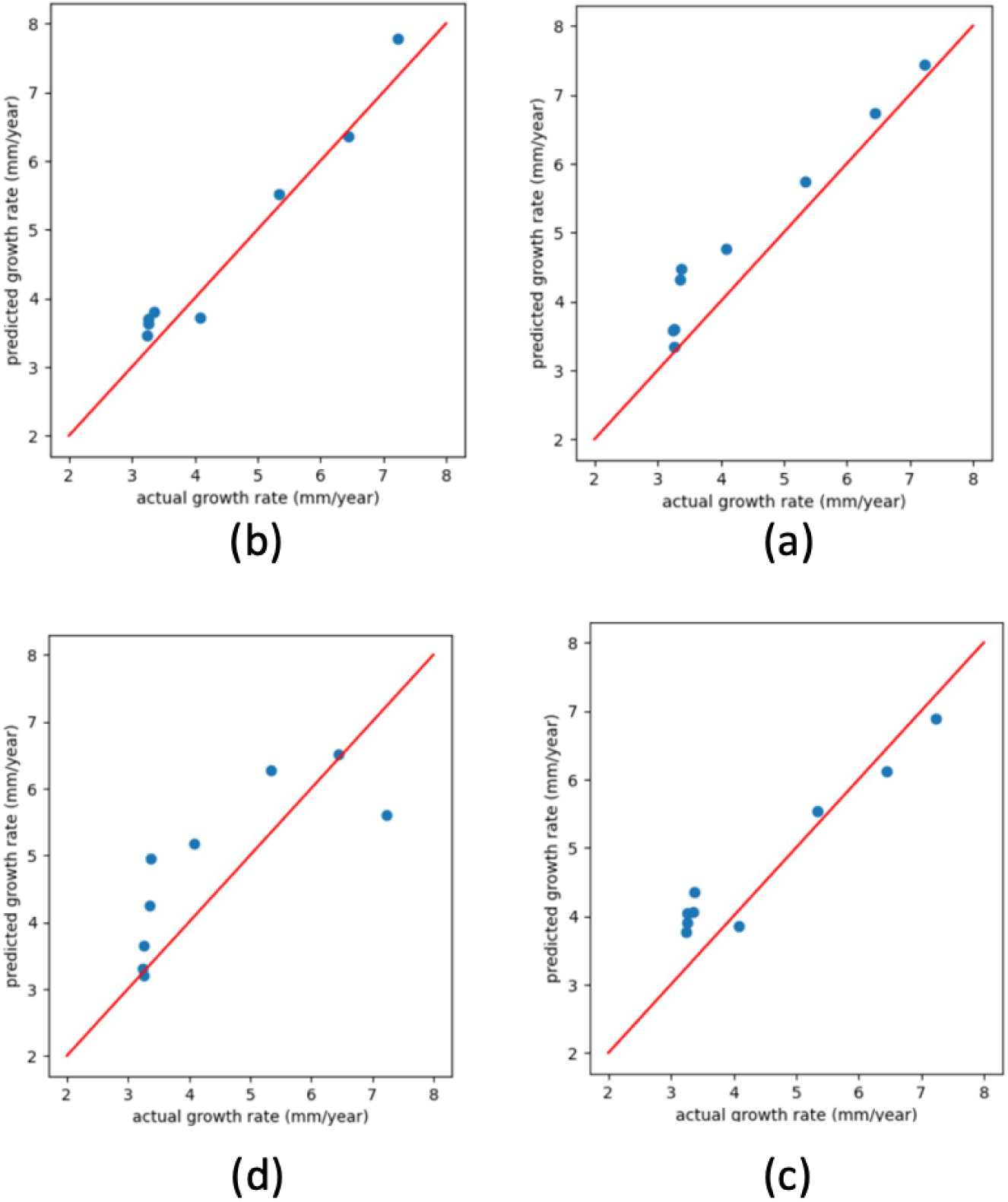
Predicted growth rate versus actual growth rate (patient data) by models a: DBN, b: RNN, c: LSTM, d: GRU.

**Table 6** evaluates the predictive performance of GRU, LSTM, RNN, and DBN for estimating AAA growth rate using RMSE and R² metrics. The RNN model performed best, achieving the lowest RMSE (0.46) and highest R² score (0.89), indicating high accuracy and reliability. DBN followed closely with an R² score of 0.86 and RMSE of 0.57, while LSTM showed competitive performance (RMSE: 0.62, R²: 0.84). GRU demonstrated the weakest predictive capability, with the highest RMSE (0.91) and lowest R² score (0.76). These results highlight RNN and DBN as the most effective models for predicting AAA growth rates.

**Table 6.** R² score and RSME for different models in predicting growth rate (patient data).

|  | GPU | LSTM | RNN | DBN |
| --- | --- | --- | --- | --- |
| RMSE | 0.91 | 0.62 | 0.46 | 0.57 |
| $R^2$ score | 0.76 | 0.84 | 0.89 | 0.86 |

**Table 7** presents the R² scores for recurrent models (GRU, LSTM, and RNN) in predicting aneurysm growth rates across different sequence lengths (one, two, and three sequences). The results reveal that RNN consistently achieved the highest R² scores across all sequence lengths, with 0.89 for three sequences, 0.83 for two sequences, and 0.77 for one sequence, indicating superior performance and a stronger ability to explain variance in growth rate predictions. LSTM also demonstrated competitive performance, with R² scores of 0.84, 0.79, and 0.73 for three, two, and one sequences, respectively, highlighting its reliability across varying input lengths. GRU, however, exhibited the lowest R² scores in all cases, achieving 0.76 for three sequences, 0.68 for two sequences, and 0.64 for one sequence, suggesting reduced predictive accuracy compared to LSTM and RNN. Overall, the results emphasize the importance of sequence length, with longer sequences improving prediction accuracy, and further underscore the RNN model’s robustness in AAA growth rate forecasting.

**Table 7.** R² score of recurrent models in predicting growth rate for different sequence lengths.

| $R^2$ score | GRU | LSTM | RNN |
| --- | --- | --- | --- |
| Three sequence | 0.76 | 0.84 | 0.89 |
| Two sequence | 0.68 | 0.79 | 0.83 |
| One sequence | 0.64 | 0.73 | 0.77 |

## 4. Discussion

In recent years, a considerable number of studies have been done on the development process of AAAs and the effect of geometric and non-geometric parameters [2,52,53]. Also, computational models of G&R have shown strong potential for advancing the treatment of arterial diseases and for improving diagnosis and outcome prediction in medicine [22,38,54,55]. Furthermore, there has been growing interest in the role of biomechanical factors affecting the aortic wall’s stress distribution and growth progression, which could have implications for improving G&R predictions [5,56,57]. Nevertheless, the application of G&R models is hampered by the difficulty of obtaining sufficient patient data. In particular, small AAAs are typically assessed using ultrasound imaging, which has limited spatial resolution, while the use of CT for longitudinal follow-up is constrained by concerns about radiation exposure [58]. Therefore, the generation of artificial (synthetic data) holds high potential for training artificial intelligence (AI) models and enhancing predictive capabilities.

This study presents an integrated framework combining a physics-based G&R model with patient-specific clinical imaging data to predict the temporal evolution of AAAs in three dimensions. To our knowledge, this is one of the first studies utilizing sequential deep learning models—RNN, LSTM, GRU—alongside a DBN, trained via a two-stage process leveraging a large, diverse *in silico* dataset augmented with clinical images, to directly predict both maximum AAA diameter and growth rate. Deep learning approaches utilizing longitudinal data have recently been applied to aneurysm expansion prediction, demonstrating their utility in personalized G&R models [3,19]. In this research, a workflow was applied, wherein the machine learning models are pre-trained with a virtually generated large-scale dataset, enhanced by surrogate modeling via kriging, and subsequently fine-tuned using real patient data. This two-stage training methodology leverages the synthetic diversity of simulated data with the realism of patient images and outperforms traditional DBN-based approaches [59]. This approach demonstrates that deep learning, when combined with data-efficient physical modeling, is well-suited for capturing the evolutionary geometry of vascular diseases and for supporting clinical decision-making.

When evaluating degradation functions for the formation and progression of AAAs, key considerations include simplicity, empirical accuracy, ability to capture biological phenomena, and computational efficiency. These functions are crucial for understanding disease progression by simulating the weakening of the arterial wall. They also integrate with G&R models to predict morphological changes, such as aneurysm growth and geometry, by modeling the balance between material degradation and synthesis. Additionally, degradation functions capture biomechanical dynamics by accounting for mechanical factors, such as blood pressure and wall stress, which influence tissue weakening and aneurysm expansion. Our simulation results explain the complex relationship between elastin degradation, increased wall stress, and compensatory collagen production, showcasing AAA growth over 3600 days. The degradation function (Eq. 1) used in our study demonstrates several strengths in modeling AAA formation and progression. It is specifically designed to improve upon traditional approaches by capturing the localized and asymmetric weakening of the aortic wall. This function incorporates a combination of two 2D Gaussian distributions, allowing it to model spatially varying elastin degradation along the axial and circumferential directions, which better reflects the heterogeneous nature of AAA growth observed in clinical data. By integrating this function with the G&R model, this approach effectively simulates morphological changes like asymmetry, tortuosity, and maximum diameter growth over time. Importantly, it accounts for the balance between material degradation and synthesis, enabling realistic simulations of AAA expansion. **Figure 5** illustrates the diverse geometries produced through simulations using the degradation function proposed (Eq. 1), capturing asymmetry, tortuosity, and realistic growth patterns seen in AAAs, which are aligned with previous studies [32]. Increased asymmetry has been associated with higher wall stress and elevated rupture risk, even in aneurysms below traditional diameter thresholds. Also, tortuosity values within the simulated range correspond to those reported in clinical cohorts and are relevant for both rupture assessment and repair planning. By reproducing these clinically observed shape characteristics, the model provides geometries that can directly support patient-specific risk evaluation and treatment decision-making.

Beyond the methodological integration of G&R simulation, surrogate modeling, and deep learning, this work also contributes in terms of computational and predictive efficiency. The surrogate-based kriging method enabled rapid synthesis of thousands of distinct AAA geometries, significantly reducing computational costs compared to traditional G&R simulations, as demonstrated by Jiang et al. [20]. The kriging approximation drastically accelerated the process, allowing for large-scale data generation suitable for deep model training, which highlights efficiency improvements over traditional G&R models while maintaining an acceptable level of approximation uncertainty [12,51]. Inspired by multiphysics features and hemodynamic modeling approaches, we believe integrating surrogate modeling can further refine predictions of AAA growth under varying biomechanical conditions [5,60].

The results in **Table 5** and **7** show that sequential architectures performed best overall: the LSTM achieved the highest accuracy for maximum diameter prediction (R² = 0.92), while the RNN provided the strongest performance for growth-rate prediction (R² = 0.89) Sequential models outperform DBNs by better capturing temporal relationships in data and addressing gradient issues, resulting in higher predictive accuracy and greater robustness for modeling complex AAA progression. **Figures 13** and **14** further validate the temporal efficacy of these models by displaying predicted growth curves closely matching real patient data from 25 clinical cases. In addition to higher accuracy, another advantage of recurrent networks is that these models have the ability to predict with variable input size. In fact, it is possible to use any number of arbitrary sequences to predict the next sequence. Therefore, in the use of this model, there will be more flexibility in choosing the input of the model, but it is obvious that the more input history, the more accurate the models will be.

Although this study advances AAA growth prediction towards better alignment with clinical data, several limitations remain that affect the current framework. Due to inconsistent timing in patient CT imaging data, interpolation was necessary to standardize time points for model training and testing, a process that may reduce flexibility and introduce bias. Additionally, although the surrogate-augmented dataset captures some aspects of morphological complexity (e.g., tortuosity), the absence of other important biochemical or anatomical variables still limits the model’s applicability across a broader range of cases. Moreover, the lack of real patient-specific initial geometries and non-morphological data, including genetic, environmental, and demographic factors such as gender, smoking history, and family history, restricts the model’s ability to fully capture individual variability. Recent clinical studies have also indicated that quantitative analysis of intraluminal thrombus (ILT) geometry and texture can provide additional predictive value for AAA growth. The absence of such ILT-specific imaging biomarkers in the current framework may therefore limit its ability to capture all relevant structural determinants of aneurysm progression [61]. Lastly, several hemodynamic variables such as low wall shear stress, high relative residence time (RRT), and complex vortical structures have been suggested to be associated with localized aortic growth, which is in turn linked to intraluminal thrombus (ILT) accumulation [55,62–64].

Therefore, future work will focus on expanding the diversity of simulated data, incorporating more patient scans with varied temporal patterns, and ultimately integrating more comprehensive models of AAA evolution, such as fluid-solid-growth frameworks [55,65–67], that combine wall stress, hemodynamics, and tissue remodeling. Additionally, with larger datasets, deeper neural network architectures can be tested and optimized for improved predictive power. Future work could also integrate G&R simulations with implicit neural representations (INRs), which can reconstruct smooth, high-fidelity AAA surfaces from minimal imaging data, thereby improving the accuracy of growth predictions [68]. We anticipate that continued refinement of both physical and data-driven models will further close the gap between in silico prediction and clinical reality, offering tangible benefits for patient monitoring and intervention planning. In summary, our integrated framework demonstrates that combining synthetic G&R-based datasets, surrogate modeling, and advanced deep learning approaches can yield highly accurate, efficient, and clinically relevant predictions of AAA growth. Our model’s performance underscores its potential for practical implementation in monitoring and risk assessment of AAAs. This research establishes a strong foundation for future studies aiming to personalize AAA management with transparency and reliability, particularly in supporting critical surgical decisions.

## Data Availability

All data produced in the present study are available upon reasonable request to the authors

## Acknowledgement

This work has been supported in part by the National Institutes of Health (1R21HL113857).

## Appendix

## A. Stress-mediated Growth and Remodeling (G&R) Model

The central concepts of the constrained mixture model (CMM) for growth and remodeling (G&R) of soft tissues were introduced by Humphrey and Rajagopal [26]. In this framework, tissue behavior is described in a homogenized sense, wherein the motion of each constituent is constrained to be the same as that of the solid mixture. This assumption neglects moment exchange among the solid constituents and thermodynamic dissipation over short time scales. However, biological tissue adaptation is governed by the continual turnover of its constituents through continual production and removal which can effectively tract long-term changes in tissue shape and structure.

The model comprises the stored energy functions *w*_*R*_(*t*; *s*) with stress-mediated growth and remodeling functions 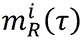, which are summarized below [22]:

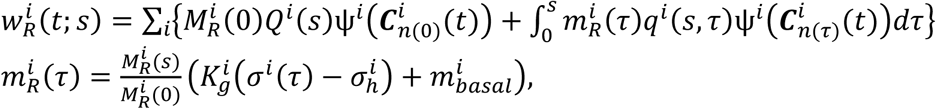

where *i* = *e*, *k*, *m* where *e* denotes elastin, *k* denotes the *k*th family of collagen fibers, and *m* denotes the smooth muscle cells; *t*, *s*, and *τ* are the current time, time in long-term scale, and the production time, respectively; 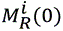 and 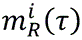 are the total area mass density at time *0* and the mass production rate of constituent *i* in the reference configuration, respectively; *Q*^*i*^(*s*) and *q*^*i*^(*s*, *τ*) are the fraction of the constituent *i* that was presented at time *0* and its survival rate function, respectively; 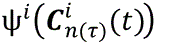 is the store energy function per unit mass which is a function of the right Green stretch tensor from the stress-free configuration of constituent *i* to the current configuration where 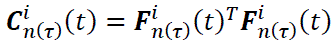 and 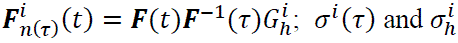 are a scalar-value of stress for constituent *i* at the production time *τ* and the target or homeostatic stress value, respectively; 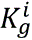 is the scalar parameter that controls the stress-mediated growth; and 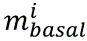 is a basal rate of mass production for constituent *i*. For the stress-mediated stress G&R model, it is assumed that elastin *e* is not produced or removed during the growth and only collagen fibers *k* and smooth muscle cells *m* are assumed to be produced depending on the stress at the production time *τ*. The rationale of G&R AAA model can be found in [37, 38, 22]

The stored energy functions for the elastin-dominated amorphous, collagen fiber families, and passive smooth muscle cells are given as

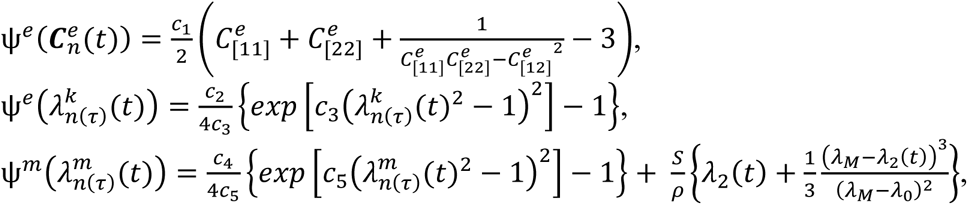

where 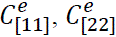, and 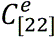 are components of 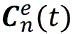 which represents the Green tensor of elastin mapping from its stress-free natural configuration to the current configuration; 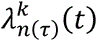 is the stretch of collagen fiber mapping from the natural stress-free configuration to the current configuration given by

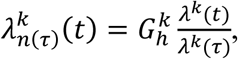

where *λ*^*k*^(*t*) and *λ*^*k*^(*τ*) denote stretch of collagen fiber mapping from the natural to the current configuration at time *t* and *τ*, respectively. Similarly, 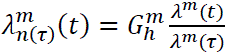. *λ*_*M*_ and *λ*_0_ represents the maximum and minimum stretch values of smooth active tone and *S* denotes the maximum stress caused by active tone.

Using a biaxial stretching test dataset provided by Geest et al. [69], the passive constitutive parameters of CMM (*c*_1_, *c*_2_, *c*_3_, *c*_4_, *c*_5_) as well as the angle of diagonal fiber family *ɑ*^*k*^ and elastin mass fraction *φ*^*e*^ were estimated via nonlinear regression for 17 subjects [70]. The resulting parameter set was used in the G&R finite element (FE) simulation.

## B. Inverse Optimization to Minimize the Deviation of Homeostatic Stress Values

Because the initial healthy aortic geometry was obtained from a medical CT image, the model must be prestressed with the in vivo blood pressure. Within the constrained mixture framework, the reference configuration is assumed to correspond to a homeostatic state, in which new constituents are deposited with a prescribed prestretch (also referred to as the deposition stretch, 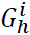). Accordingly, the imaged geometry is assumed to represent a homeostatic configuration under the mean in vivo blood pressure. However, because patient-specific geometries derived from medical images are inherently curved and heterogeneous, the stress distribution under the mean arterial pressure deviates from the target homeostatic values. To address this discrepancy, an inverse optimization approach was employed to adjust vessel wall thickness and collagen fiber orientation such that the computed stresses match the homeostatic target values throughout the geometry [33,34].

The spatial distribution of wall thickness *h* and the orientation of diagonal fiber family *ɑ*^*k*^, treated as an independent variable governing material anisotropy, using an inverse optimization approach that minimizes both deviations of geometry from the in vivo configuration and deviations of stress from the target homeostatic values. The objective function to minimize is

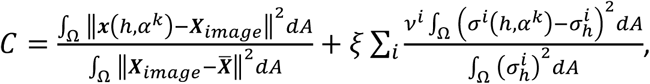

where *i* = *m*, 1, …, *k*. ***x*** is the FE simulation for position vector, ***X***_*image*_ is the position vector from medical image, and ***̅X*** is the geometric center of the artery. *σ*^*i*^is a scalar measure of stress in the direction of constituent *i* obtained from the FE analysis. The set (ℎ, *ɑ*^*k*^) are the unknown variables to be minimized with a weight parameter *ξ*. However, solving the optimization problem with geometrical variables at all nodal points of the FE model would incur excessive computational cost. Instead, the spatial distributions of wall thickness and anisotropy were approximated using a small number of variables with associated basis functions, defined independently of the FE mesh as

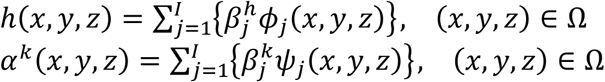

where 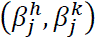 are variables for thickness and anisotropy associated with the approximation point *j*. *φ*_*j*_(*x*, *y*, *z*) and *ψ*_*j*_(*x*, *y*, *z*) are basis/approximation functions defined on the computational domain Ω. The Nelder-Mead Simplex method is used for the optimization

**Figure A1.**
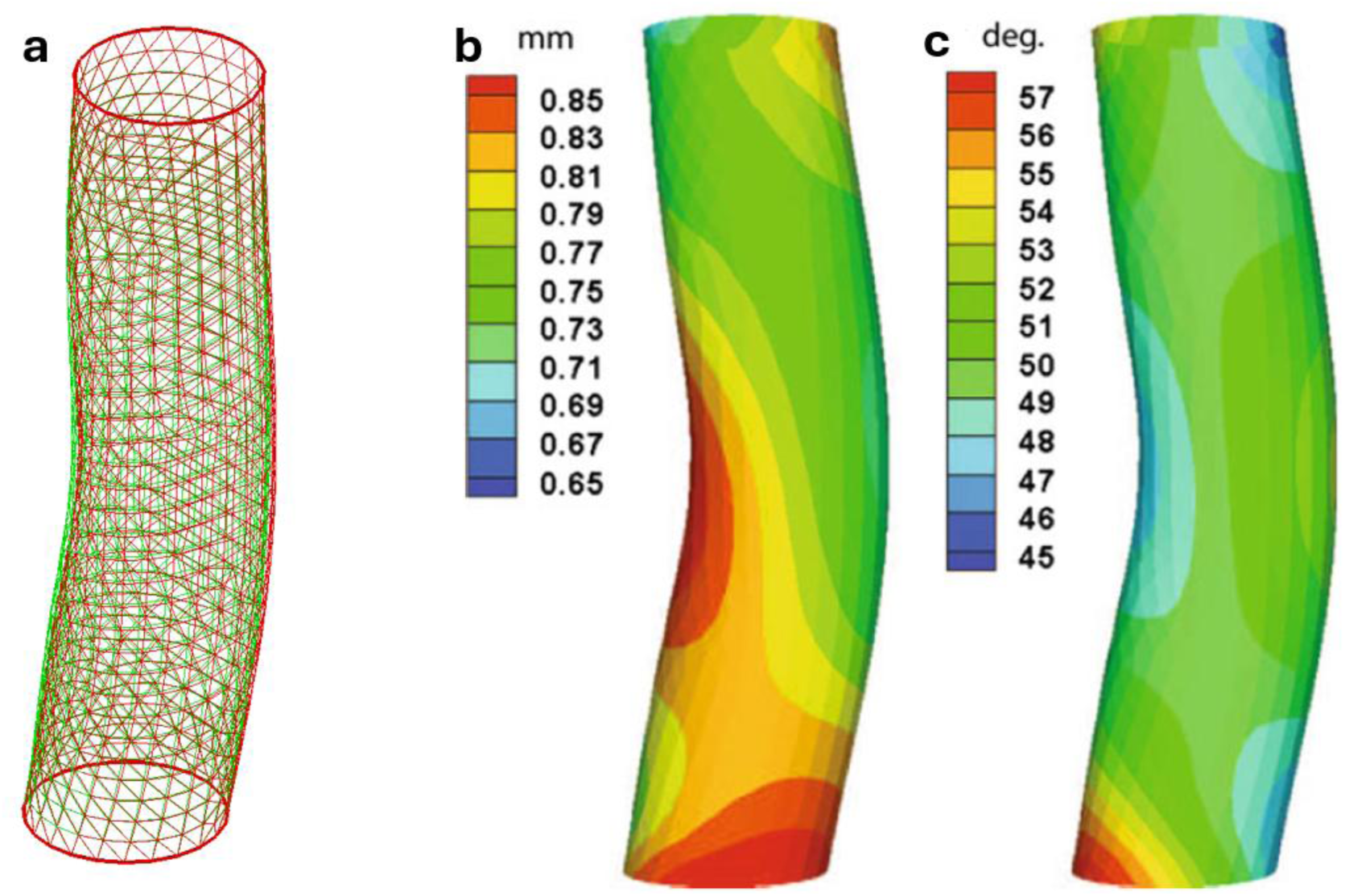
The optimization results illustrate: (a) a comparison between the original aortic wall mesh and the optimized geometry at the homeostatic state; (b) the optimized wall thickness distribution; and (c) the orientation of the diagonal collagen fiber family.

## C. Finite Element Formulation

The G&R model was originally implemented using custom MATLAB scripts but has since been rewritten in FEniCS. Cross-verification between the two implementations was performed for growth simulations (**Fig. A2**), and the FEniCS-based model was subsequently used for multi-fidelity surrogate G&R modeling to predict AAA growth [20].

The principle of virtual work is formulated for the weak form of the stored energy:

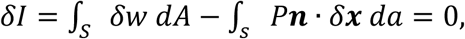

where *P****n*** denotes the inner pressure vector applied to a vessel wall; *S* and *s* correspond to the surface of the blood vessel in the reference and current configurations, respectively; *δ****x*** denotes the virtual changes in positions. The current location is replaced by FE approximation, i.e.,

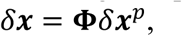

where ***x***^*p*^ is the nodal vector for the current position and ***ɸ*** is the shape function matrix. The governing equation defined on nodal points of local element is formulated by

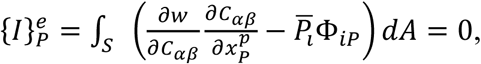

where 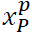 is the index notation of current position vector ***x***^*p*^, and *̅P*_*i*_ is given by

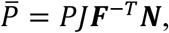

where ***N*** is the normal vector and *J* is the Jacobian. The governing equation is solved by the Newton-Raphson method.

**Figure A2.**
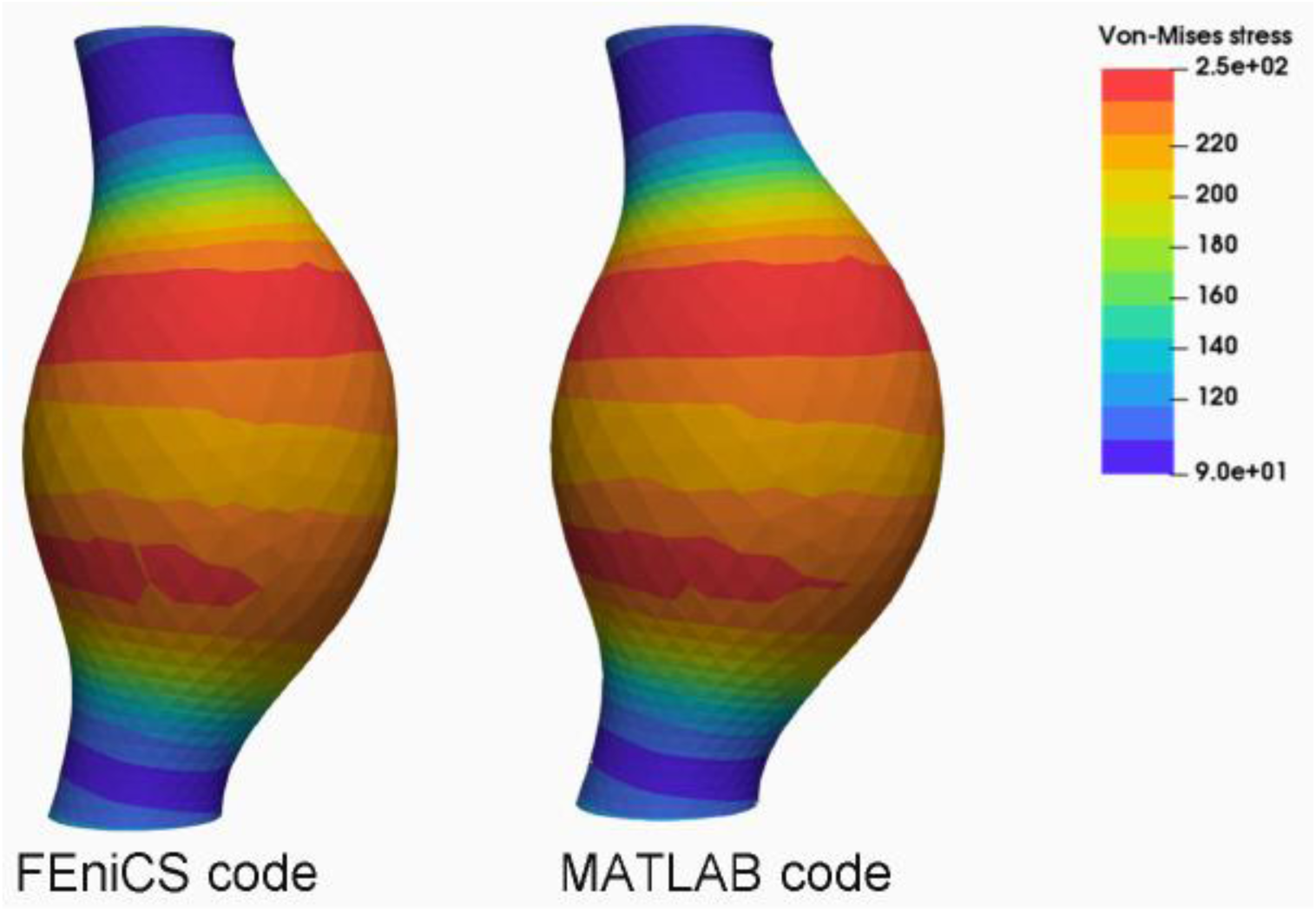
A verification between the 3D membrane G&R simulation implemented from FEniCS and Matlab, respectively.

